# Cost-effectiveness of Ultrasound Screening for Uterine Fibroids in the United States

**DOI:** 10.64898/2026.03.10.26347936

**Authors:** Pauras Mhatre, Lily von Rosenvinge, Akanksha Suresh, Kristin Patzkowsky, Anja Frost, Maria Victoria Vargas, Harold Wu, Karen Wang, Khara Simpson, James H. Segars, Bhuchitra Singh

## Abstract

**Background:** Uterine fibroids cause significant morbidity, psychosocial stress, and poor quality of life due to symptoms including heavy menstrual bleeding, anemia, pain, and bulk symptoms, as well as reproductive complications including infertility, early pregnancy loss, and preterm birth. Fibroids represent a 42.2 billion USD annual economic burden to the United States healthcare system. Despite reported delays in diagnosis of fibroids even in symptomatic women, clinical guidelines do not recommend screening for fibroids. High risk patient groups are well known. Earlier detection of fibroids through ultrasound screening could allow for earlier intervention with secondary prevention strategies or less invasive treatment options and improve the quality of life of women living with fibroids.

**Objective:** The study aimed to evaluate the cost-effectiveness of annual ultrasound screening for fibroids in women aged 25-54 years in the United States.

**Study Design:** In this economic evaluation, conducted in January-February 2026, a decision-analytic Markov model was developed using a healthcare payer perspective to analyze the cost-effectiveness of ultrasound screening for women in the United States. The time horizon was 25 to 55 years of age. Costs were adjusted for inflation to 2025 average according to the yearly medical care index of the United States consumer price index. Discounting (3% per cycle) and half-cycle corrections were calculated. Deterministic and probabilistic sensitivity analyses were performed to explore uncertainty, analyzed using TreeAge Pro Healthcare software. Model variables were obtained from published literature. All women residing in the United States aged 25-54 years were assumed to have been invited to the screening program.

**Results:** Ultrasound screening for fibroids for women was found to be not only cost-effective but also cost-saving, with an incremental cost-effectiveness ratio (ICER) of -$56,605.631 per QALY (quality-adjusted life-year) gained in the base-case analysis, at a willingness to pay threshold of $30,000 per QALY. Ultrasound screening was cost-effective at all starting ages from 25 to 54 years, with even greater benefit at younger ages. Sensitivity analyses demonstrated the robustness of these findings across a wide range of variable ranges. Ultrasound screening for fibroids showed a cumulative potential to save $1,169 billion and increase 20.7 million QALYs per year compared to no screening for a population of 63.89 million American women between 25 and 54 years old. The subset of 9.32 million Black American women experienced greater benefits, with potential savings of 183 billion and an increase of 3 million QALYs.

**Conclusion:** Based on the model-based analysis, annual ultrasound screening for uterine fibroids for women aged 25-54 years in the United States was cost-effective and cost-saving, even more so for Black women. These model-based findings highlighted the potential value of guidelines for annual ultrasound screening for fibroids, which could enable earlier diagnosis, secondary prevention, and timely intervention, with positive impact on both quality of life and healthcare costs.

**Tweetable Statement:** Annual ultrasound screening for uterine fibroids in U.S. for women aged 25–54 years was cost-effective and cost-saving.

**Study at a Glance:** *A. Why was this study conducted?:*

- To evaluate whether annual ultrasound screening for fibroids in women aged 25–54 years in the U.S. is cost-effective.

*B. What are the key findings?:*

- Annual ultrasound screening beginning at 25 years was both cost-effective and cost-saving, with an ICER of –$56,605.631/QALY for women in the US.
- Screening resulted in potential savings of $1,169 billion for US healthcare payers and 20.7 million QALYs for U.S. women.

*C. What does this study add to what is already known?:*

- Annual ultrasound screening for fibroids is not only cost effective but also cost saving, highlighting its potential to reduce diagnostic delays and enable earlier, less invasive interventions.
- The results support development and implementation of fibroid screening guidelines.

## Introduction

Uterine fibroids are the most common benign tumors affecting women of reproductive age,^1^ with an estimated cumulative incidence of ∼70% for white women and >80% for Black women by the age of 50 years.^2^ Fibroids contribute to significant morbidity and poor quality of life due to symptoms including heavy menstrual bleeding, anemia, pain, and bulk symptoms,^3^ as well as reproductive complications including infertility, early pregnancy loss, and preterm birth.^1,2,4–6^ Fibroid disease exhibits a rising incidence and disease burden globally,^2,7^ and living with uterine fibroids is a significant psychosocial stressor.^5^ Notably, Black women are disproportionately affected by uterine fibroids because they experience a higher prevalence of fibroids and are more likely to be diagnosed with multiple and larger fibroids.^2,8,9^ In addition to biological differences, systemic racism and inequities in care further contribute to healthcare neglect and delayed diagnosis among Black women.^1,10,11^

Reports indicate that many women with fibroids remain undiagnosed and frequently experience delays in diagnosis.^6,11^ Delays up to three to five years are reported in symptomatic women,^6^ and result in many women having to endure a significant symptom burden without a clinical diagnosis.^1,12,13^ Women may reinterpret or normalize fibroid-related symptoms, particularly non-bleeding manifestations such as pain or bulk symptoms, which may contribute to under-recognition and treatment delays.^14^ Such delays in diagnosis and treatment are major contributors to the estimated annual economic burden of $42.2 billion in the United States.^15,16^ However, despite high risk segments of the population with a disease prevalence exceeding 50%, no clinical guidelines recommend screening for fibroids. Other screening programs for diseases that have a lower incidence in the target population have demonstrated improved outcomes with favorable cost-effectiveness, such as those for lung cancer,^17^ cervical cancer,^18^ cardiovascular diseases, and diabetes.^19^ Screening helps to identify people in a general population who are at higher risk of a health problem or a condition.^20^ By identifying conditions or risk factors at an early stage of a disease, screening programs may reduce the severity of a condition and allow a wide range of options for prophylaxis and effective treatment.^21^

Ultrasonography is an economical^22^ and feasible imaging modality for detecting fibroids with its high sensitivity and specificity ^23–24^ acknowledged by the American College of Obstetricians and Gynecologists (ACOG) and American Association of Family Physicians (AAFP).^25–26^ Early detection of fibroids through ultrasound screening could allow for the use of medical therapy or minimally invasive surgery, theoretically reducing the need for more expensive and complex surgeries typically required to treat a more extensive burden of disease.^27,28^ Hence, the objective was to evaluate the cost-effectiveness of annual ultrasound screening of women aged 25-54 years in the U.S. for uterine fibroids, compared with the current practice of no ultrasound screening for uterine fibroids.

## Materials and Methods

### Study Design and Decision Tree

This cost-effectiveness evaluation was conducted in January-February 2026 in accordance with the recommendations of the Second Panel on Cost-Effectiveness in Health and Medicine and the Consolidated Health Economic Evaluation Reporting Standards 2022 (CHEERS 2022) Statement (Table A.1). A decision-analytic Markov model was developed for assessing cost-effectiveness using a U.S. healthcare payer perspective. The study analyzed two strategies, where women either received or did not receive ultrasound screening at age 25. The target population was all adults with a uterus (“women” in this article) aged 25-54 years residing in the United States. Considering the higher prevalence and earlier incidence of fibroids in Black women,^2,8,9^ a model was also run with the same variables for Black women in the U.S. The time horizon was chosen from 25 years up to 55 years of age (age of menopause including 90% population) because the risk of fibroids, their symptoms, and complications decrease considerably after menopause. A cycle length of one year (annual screening) was chosen. Based on a state-transition diagram (Figure 1), a decision tree (Appendix C) was developed analyzing two strategies, where women either received or did not receive ultrasound screening starting at age 25. In the Markov model, if a woman was diagnosed to have symptomatic fibroids, they were treated (or no treatment) based on their choice, resulting in either treatment success (recurrence-free following treatment), or failure (accounting persistence of symptomatic fibroids due to failure of treatment itself as well as for recurrence). Women with asymptomatic fibroids were not treated in the decision tree, while still allowing for the possibility that these fibroids could become symptomatic in future cycles.

**Figure 1:**
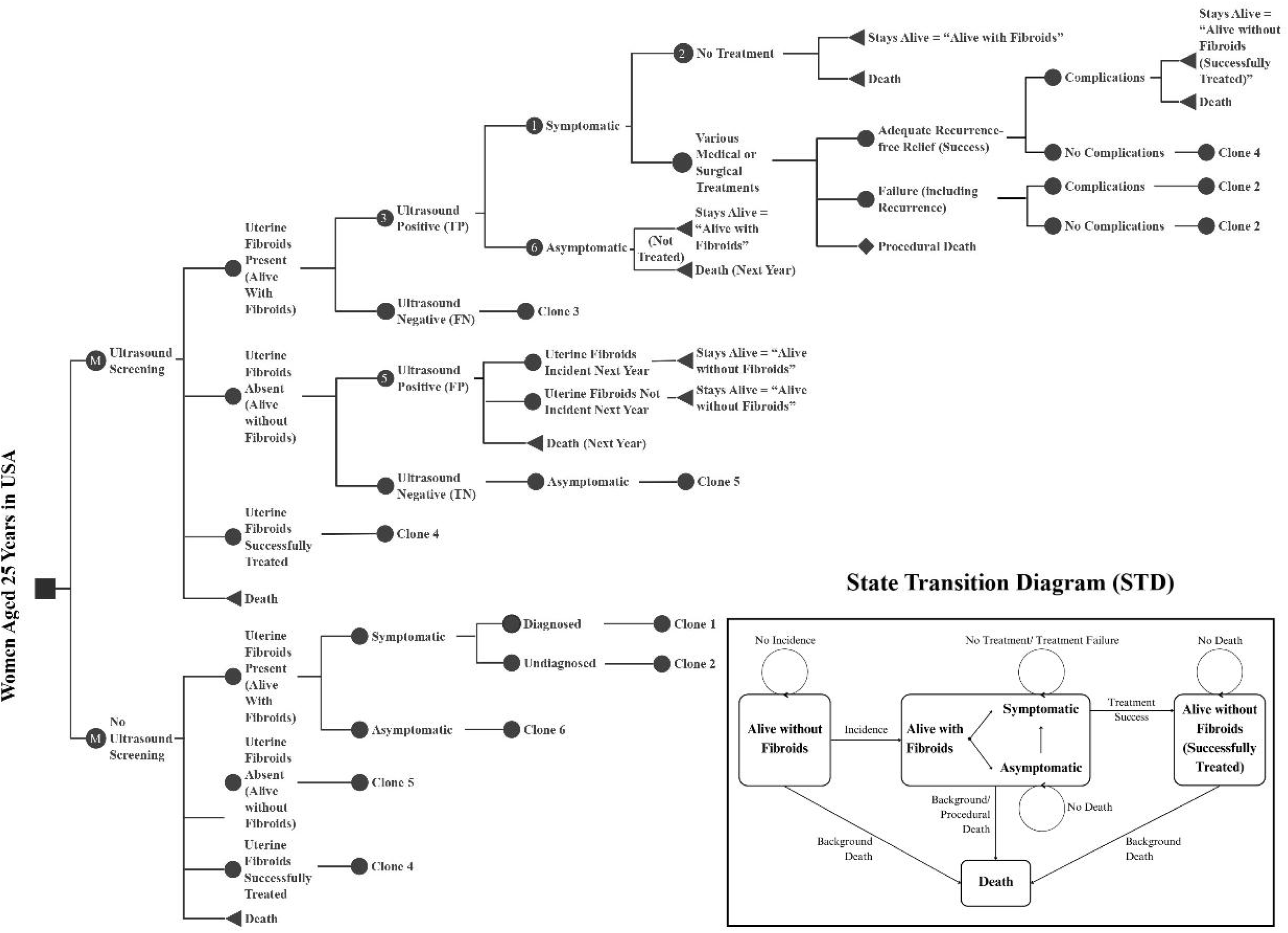
Markov Model of Ultrasound Screening for Uterine Fibroids in the United States. Health states in the model are shown in the state transition diagram which are ‘Fibroids absent’, ‘Fibroids present’, ‘Fibroids treated’, and ‘Death’. Over the course of 30 years of time horizon (25 to 54 years of age), transitions between one health state and another occurred every year, which are indicated by pointing arrowheads in the inset. ‘Women aged 25 in the U.S.’ was the ‘Decision Node’ representing two options to be compared. ‘Ultrasound screening’ and ‘No ultrasound screening’ were the Markov nodes. The diamond nodes were terminal nodes, which represented the end of the respective cycle, where transitions between health states occurred (‘jumped’ back to one of the branches of the Markov node). ‘Clone’ means that the node had similar further branches like the Master Node (numbered); where the probabilities and pay-offs (costs and utilities) might be the same or different as applicable. In the Markov model, women transitioned between four health states: Alive without Fibroids (“Uterine Fibroids Absent” arm), Alive with Fibroids (Symptomatic or Asymptomatic) (“Uterine Fibroids Present” arm), Alive without Fibroids (Successfully Treated) (“Uterine Fibroids Successfully Treated” arm), and Death. If a woman was diagnosed to have symptomatic fibroids, they were treated (or no treatment) based on their choice, resulting in (i) treatment success (recurrence-free following treatment, moving to the “Uterine Fibroids Successfully Treated” arm), (ii) failure (accounting persistence of symptomatic fibroids due to failure of treatment itself as well as for recurrence, returning to the “Uterine Fibroids Present” arm), or (iii) procedural death. Women with asymptomatic fibroids were not treated in the decision tree and were retained in the “Uterine Fibroids Present” arm, while still allowing for the possibility that these fibroids could become symptomatic in future cycles, at which point treatment decisions applied. Whereas, in the No Ultrasound Screening strategy, only the women diagnosed after being symptomatic were treated, with following branches similar to the Ultrasound Screening strategy. **Abbreviations:** TP = True Positive, FN = False Negative, FP = False Positive, TN = True Negative

### Model Variables

All variable values of the Markov model, including probabilities, costs, and utilities, are summarized in Table 1^6,16,22,23,29–59^ with ranges and distribution details (with detailed description of calculations and assumptions in Appendix B). The model variables were obtained through a thorough review of scientific literature on PubMed and Embase. Cycle-dependent variations were incorporated for prevalence, death, and treatment choices. When multiple values were available, averages were calculated; when unavailable, values were derived from indirect data or well-vetted online resources. All values were validated by six clinical specialists with expertise in benign gynecology.

**Table 1:**
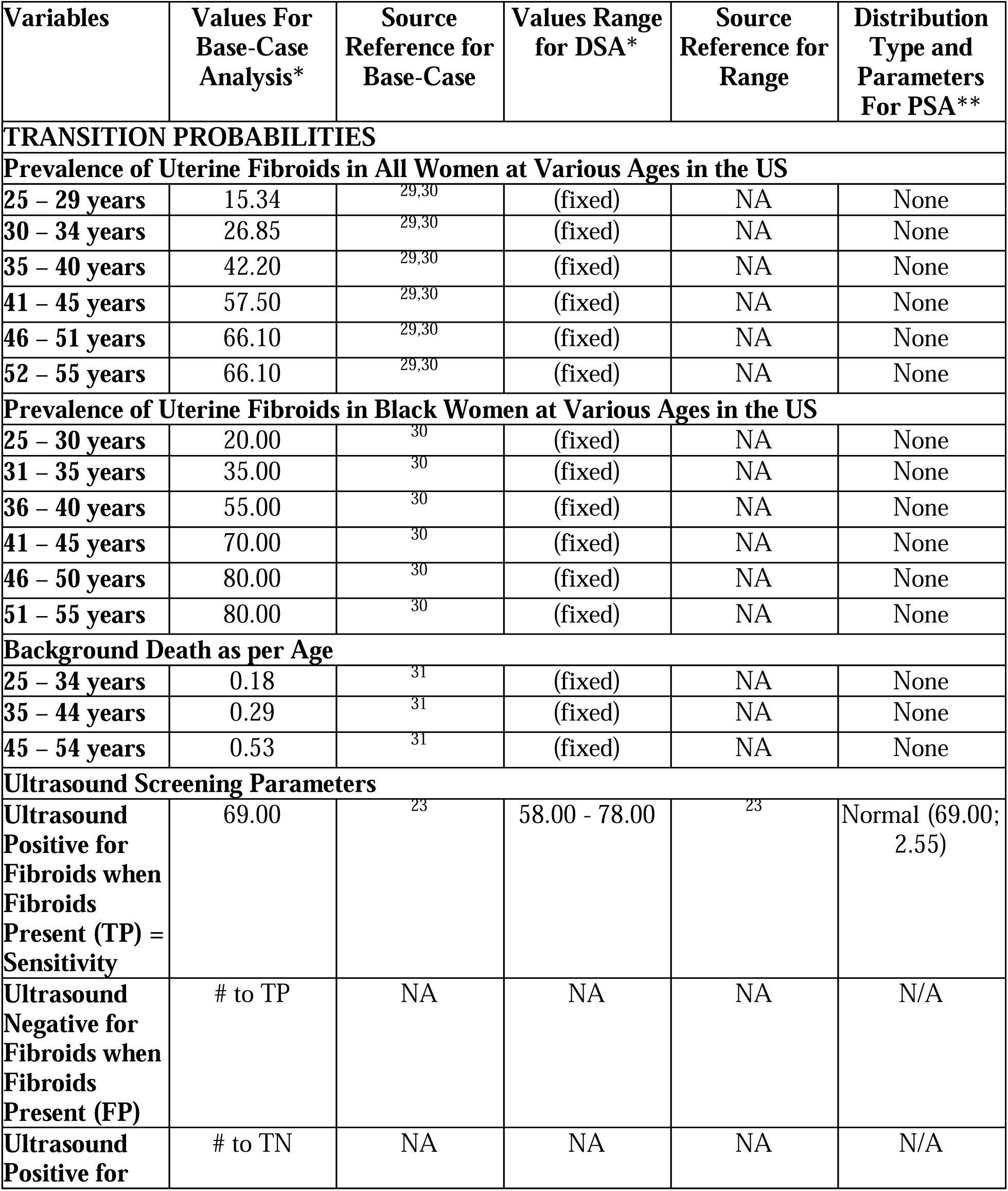
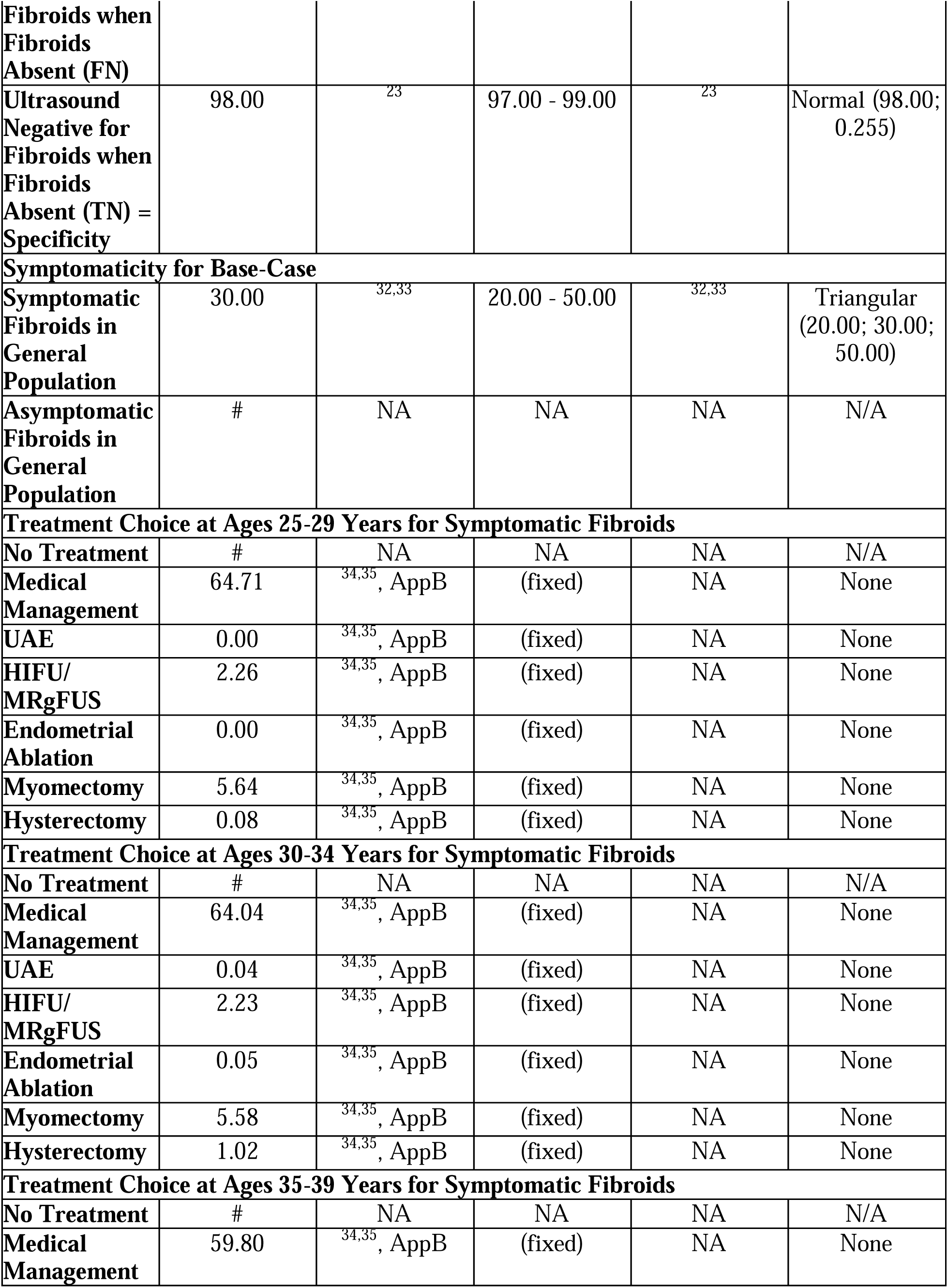
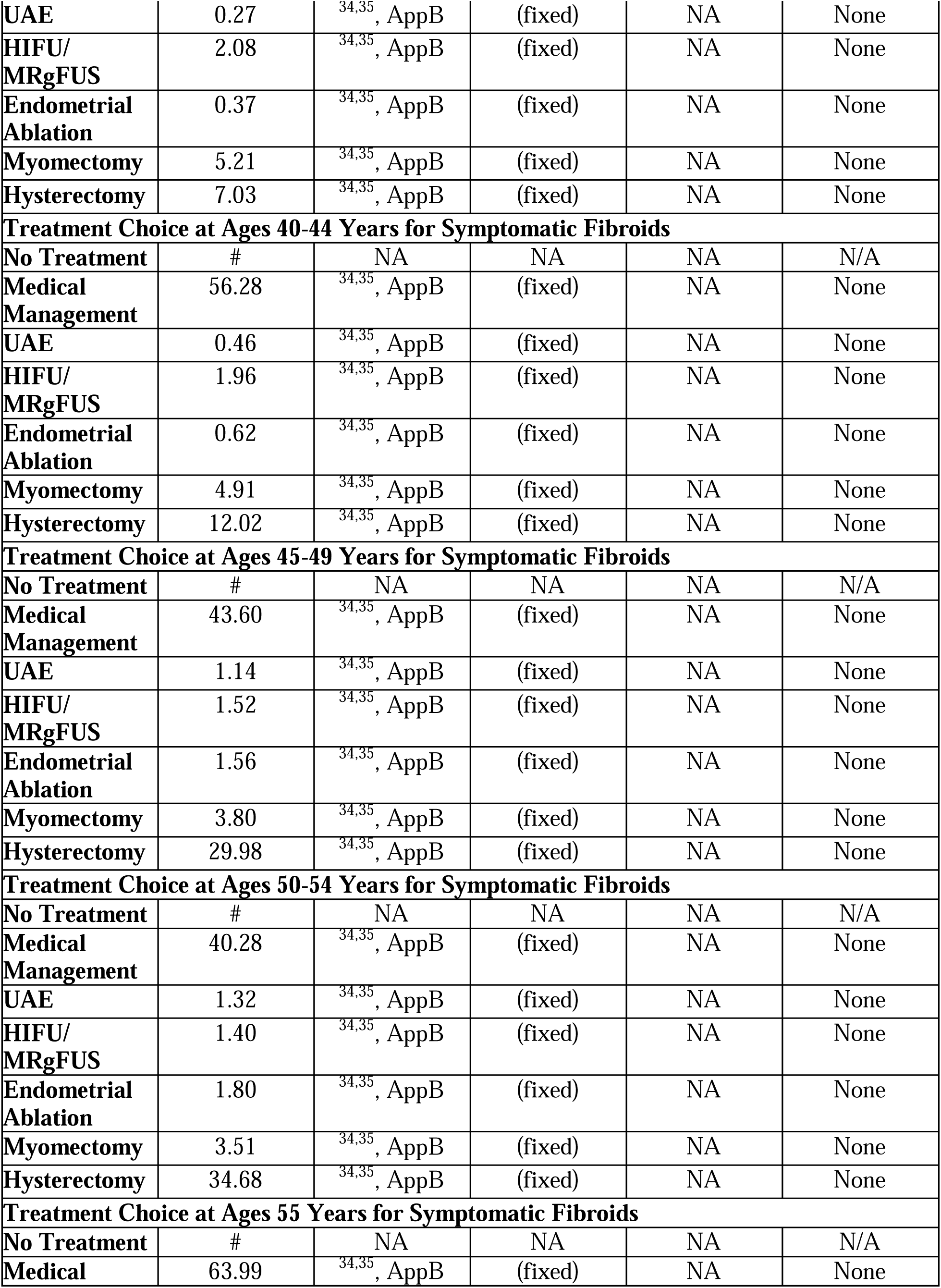
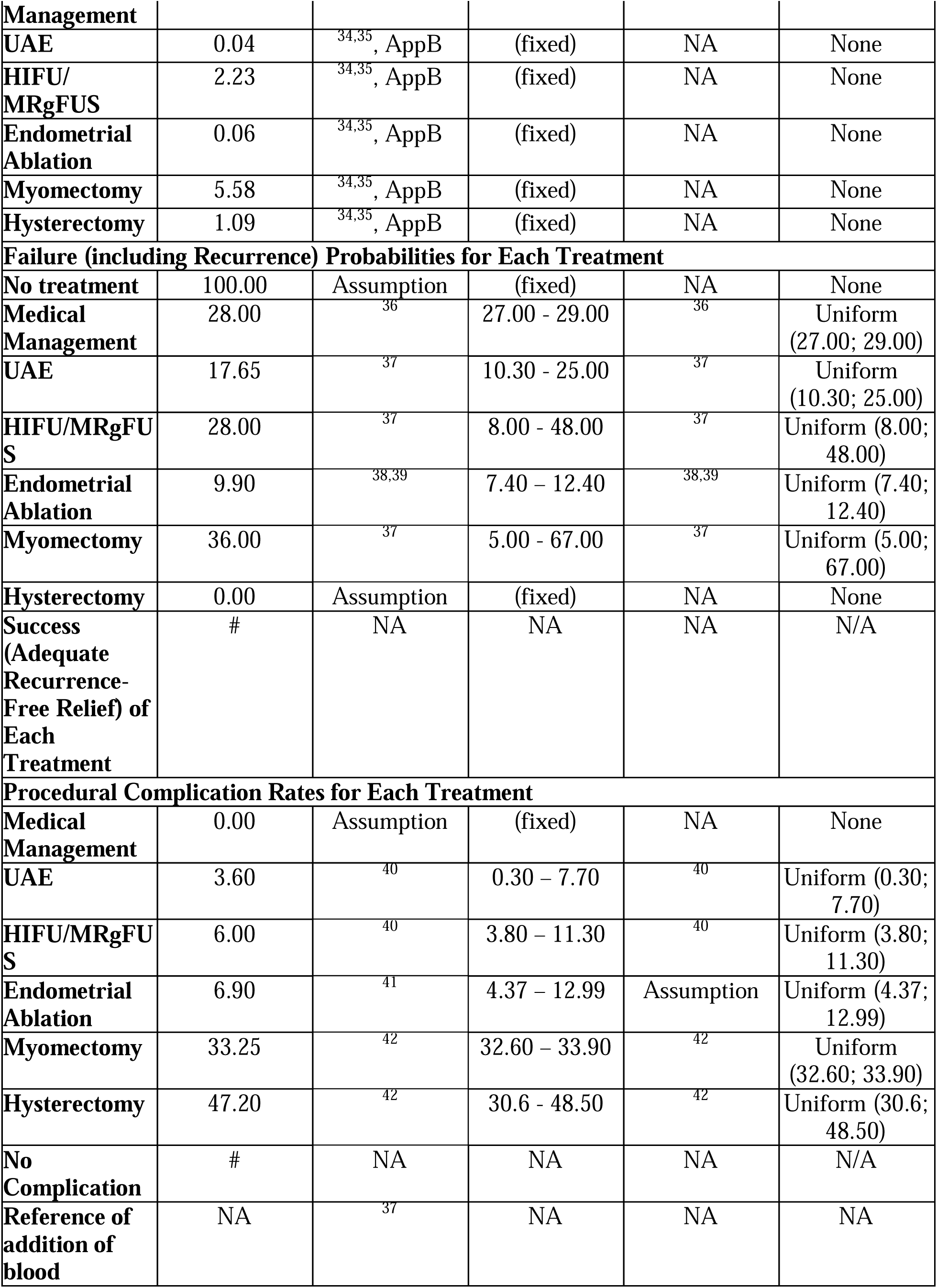
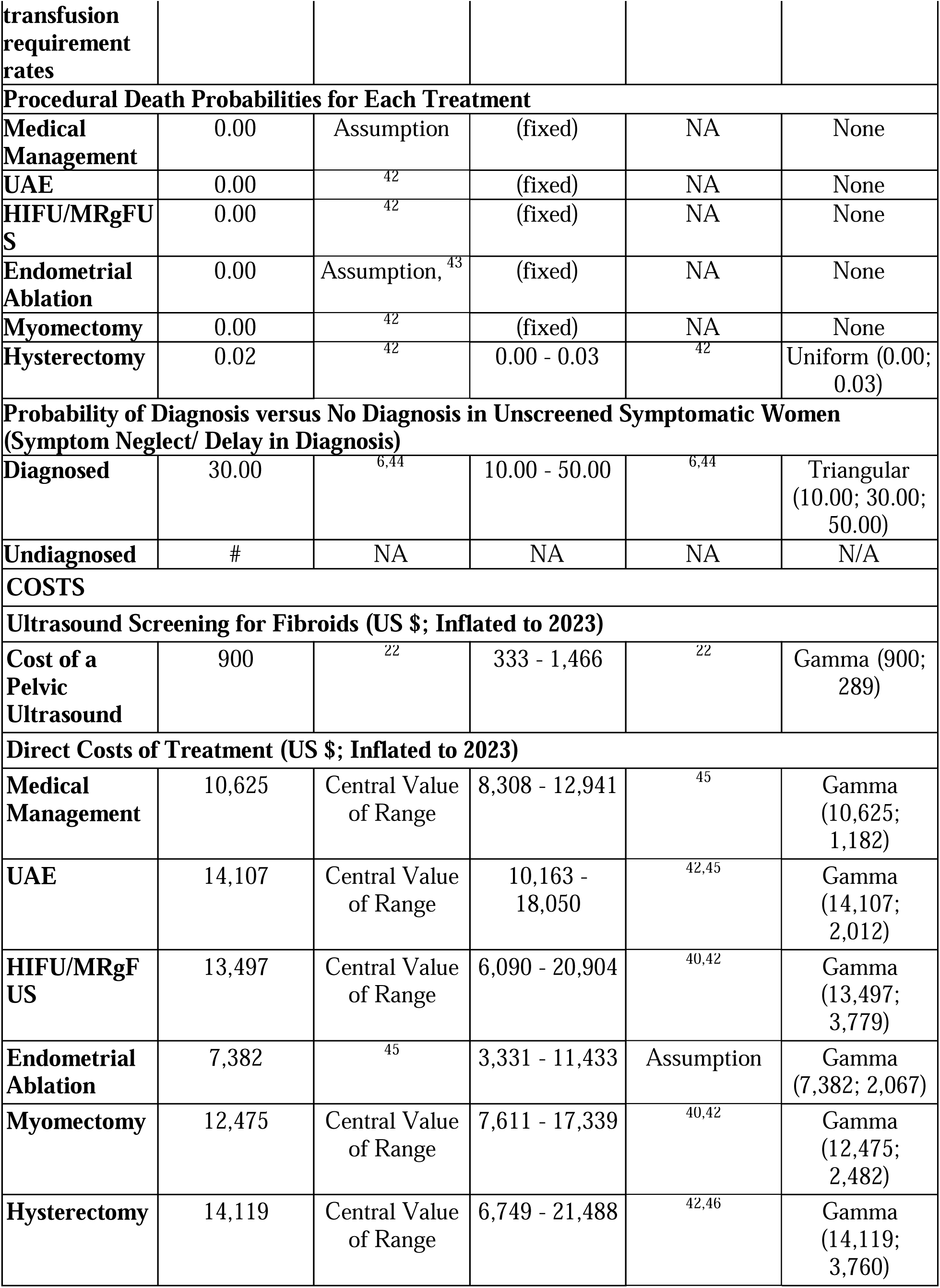
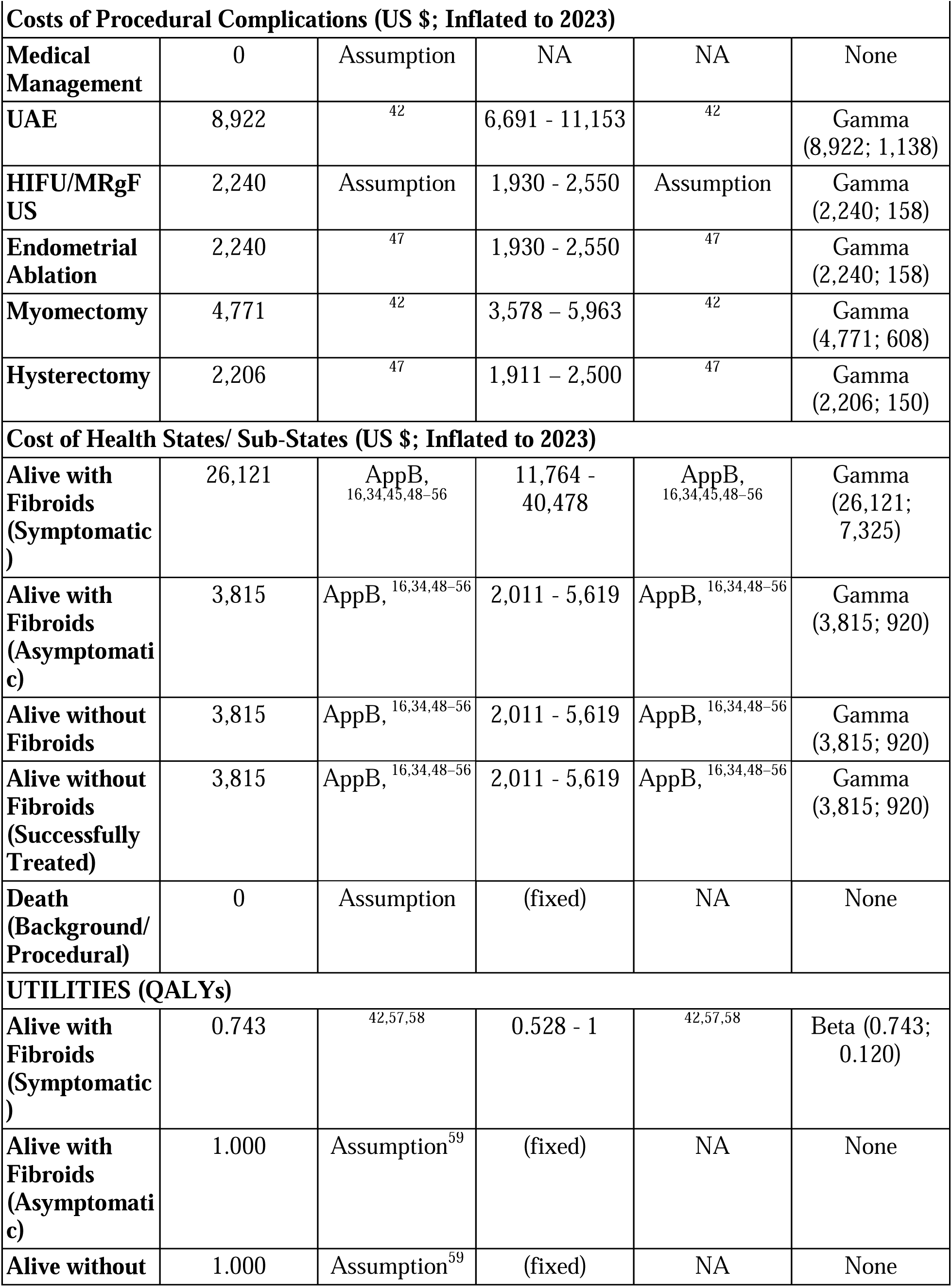
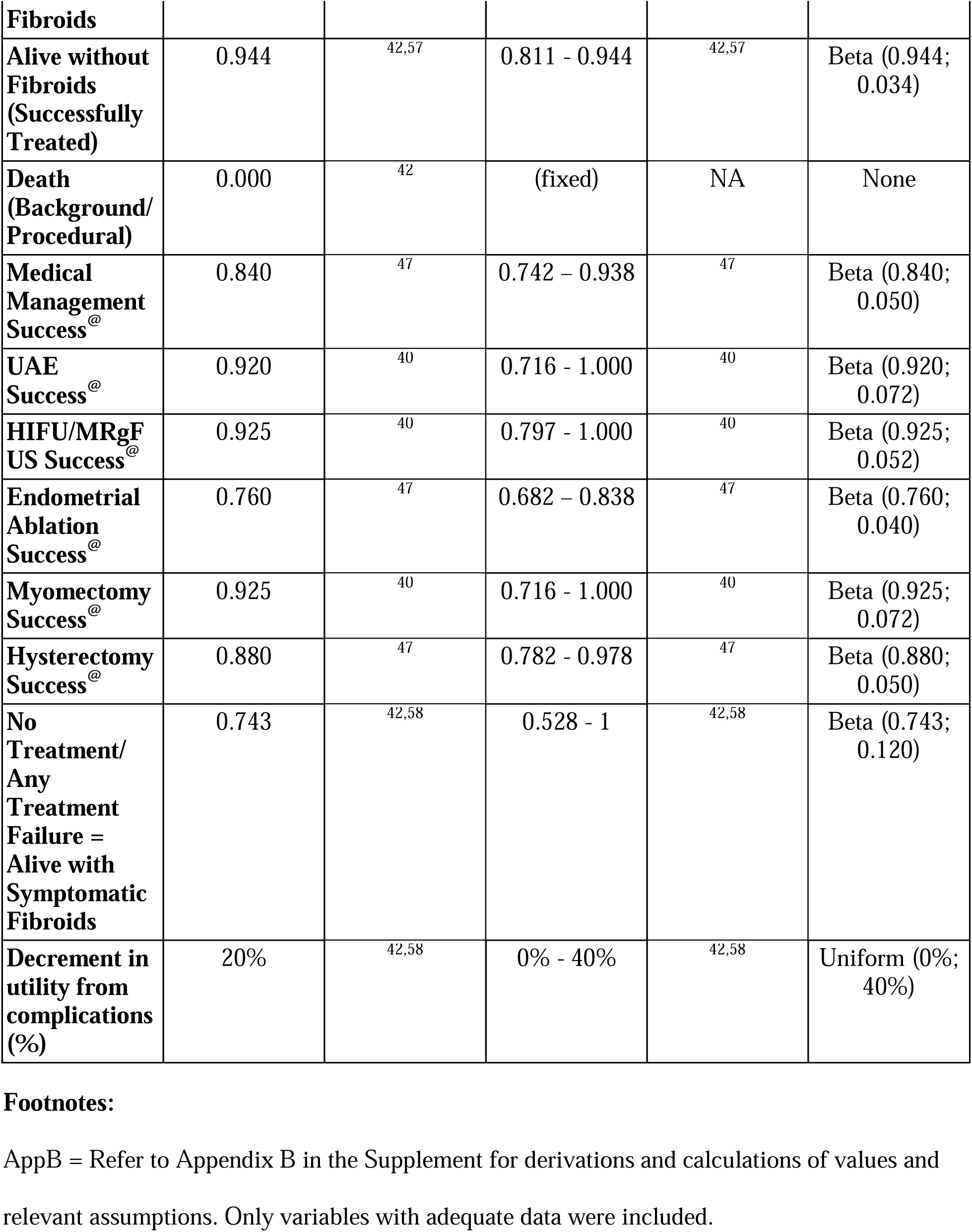
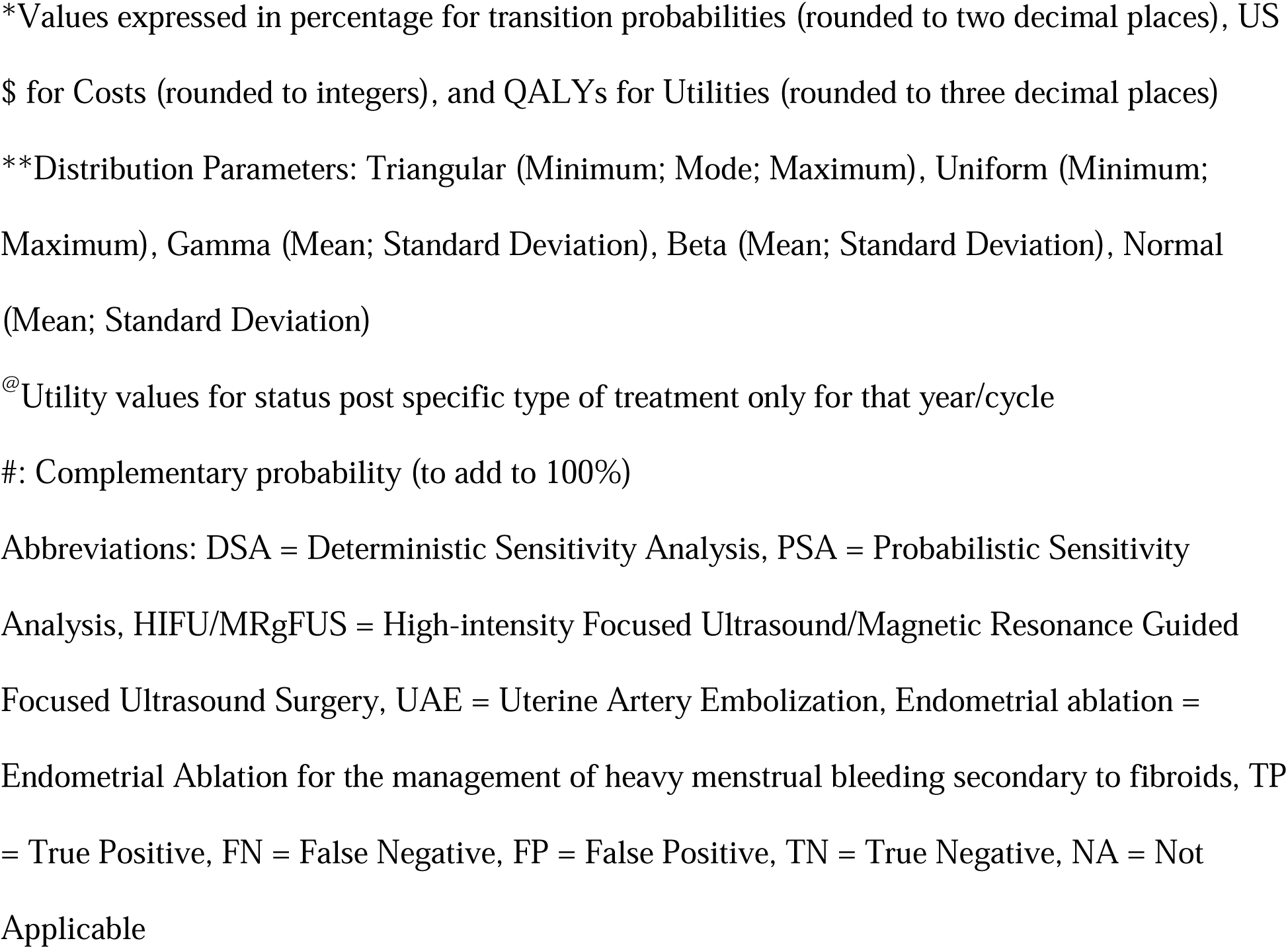
Values of Transition Probabilities and Pay-offs (Costs and Utilities) Used in the Markov Model for Cost-Effectiveness of Ultrasound Screening for Uterine Fibroids in the United States.

Probabilities reflected how women transitioned between health states. A statistical probability of death was considered at every cycle. If the patient did not receive ultrasound screening for fibroids, which is the current practice, then a woman was treated only if clinically diagnosed. Costs were based on direct costs of screening tests, interventions, and complications, inflated using the Medical Care Index of the Consumer Price Index (CPI) from annual average of the source year USD to annual average 2025 USD.^60^ Costs of living were attributed to conditions associated with fibroids, owing to a higher risk of spontaneous abortion, preterm delivery, cesarean delivery, anemia, endometriosis, inflammatory diseases, and menstruation disorders in women with symptomatic fibroids (as shown in Appendix B). The decision tree was limited to these conditions for simplicity and a conservative approach. Costs due to infertility were excluded, given that fibroids are the primary cause of infertility in about 2-3% of cases.^61^ Indirect costs associated with the loss of productivity were also excluded. Disutilities were applied to the utility of living with symptomatic fibroids per cycle for the worsening of symptoms over time (as explained in Appendix B), and to the occurrence of procedural complications when treated in a particular cycle.

### Outcomes and Analysis

All analyses were conducted using TreeAge Pro Healthcare software version 2025.^62^ The model incorporated half-cycle corrections and 3% discounting per cycle for pay-offs.^63^ The main outcomes were costs, QALYs, incremental cost-effectiveness ratios (ICERs), incremental net monetary benefits (INMBs), absolute numbers of cases detected, interventions performed, and cumulative effect on U.S. healthcare spending and QALYs. Cost-effectiveness was reported in terms of ICERs and INMBs. A conservative willingness-to-pay (WTP) threshold of $30,000/QALY was considered.^64^

A base-case analysis was conducted deterministically, assuming no uncertainty in model variables. Absolute cumulative numbers of cases detected and interventions performed over 30 years in a cohort of 10,000 women were obtained by multiplying relevant nodal probabilities by the cohort size, which represented cumulative totals over 30 years including repeated events. Case detection was estimated from true-positive ultrasound cases (screening) and diagnosed cases (no screening), while interventions were based on treatment nodes in both strategies.

Deterministic/ univariate sensitivity analyses (DSAs) were conducted using ranges to evaluate uncertainty in individual variables, represented graphically in a Tornado diagram. Alternative analyses examined the effect of age at starting ultrasound screening on outcomes. Probabilistic sensitivity analyses (PSAs) were conducted using a second-order Monte-Carlo simulation for 10,000 trials and 1,000 samples to estimate the effect of varying variables simultaneously.

## Results

In a base-case analysis for all women, ultrasound screening was found to have higher effectiveness and lower costs, making it a dominant (cost-saving) strategy. As shown in Table 2, ultrasound screening was dominant with an ICER of -$56,605.631 per QALY gained and an INMB of $28,001.314. Similar results were found in a base-case analysis for Black women with an ICER of -$60,934.859 per QALY gained and an INMB of $29,374.165 (Table 2).

**Table 2:**
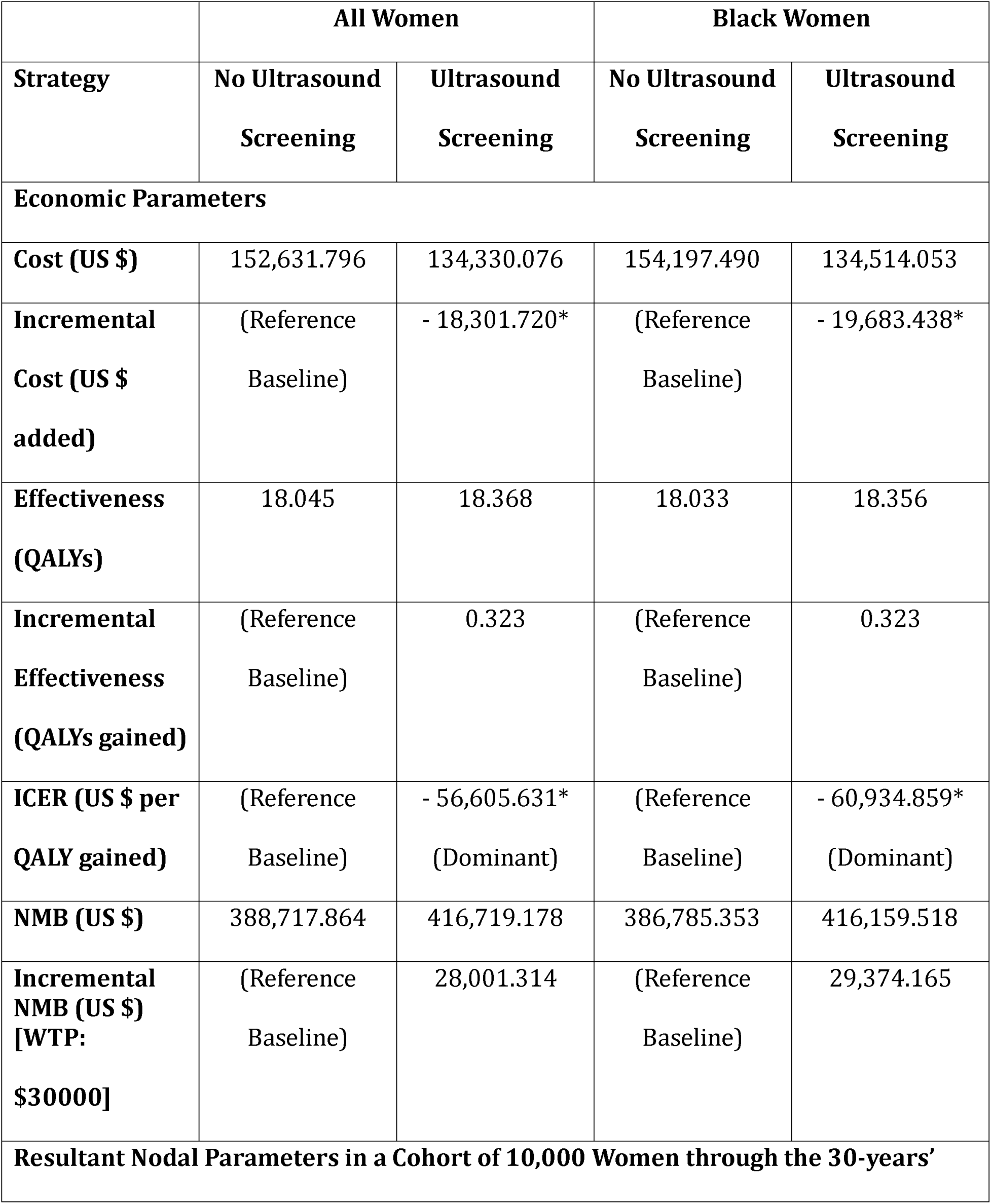
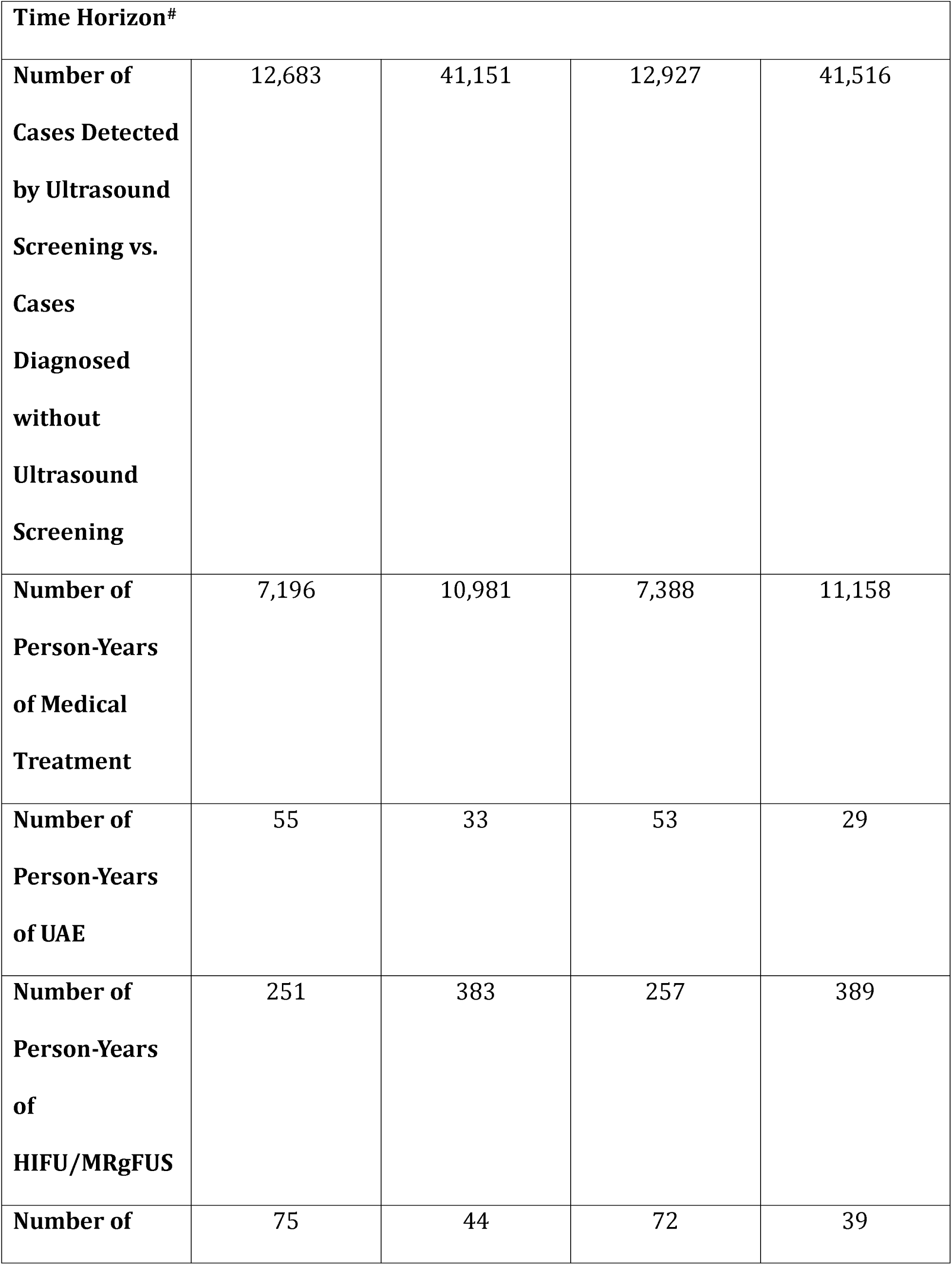
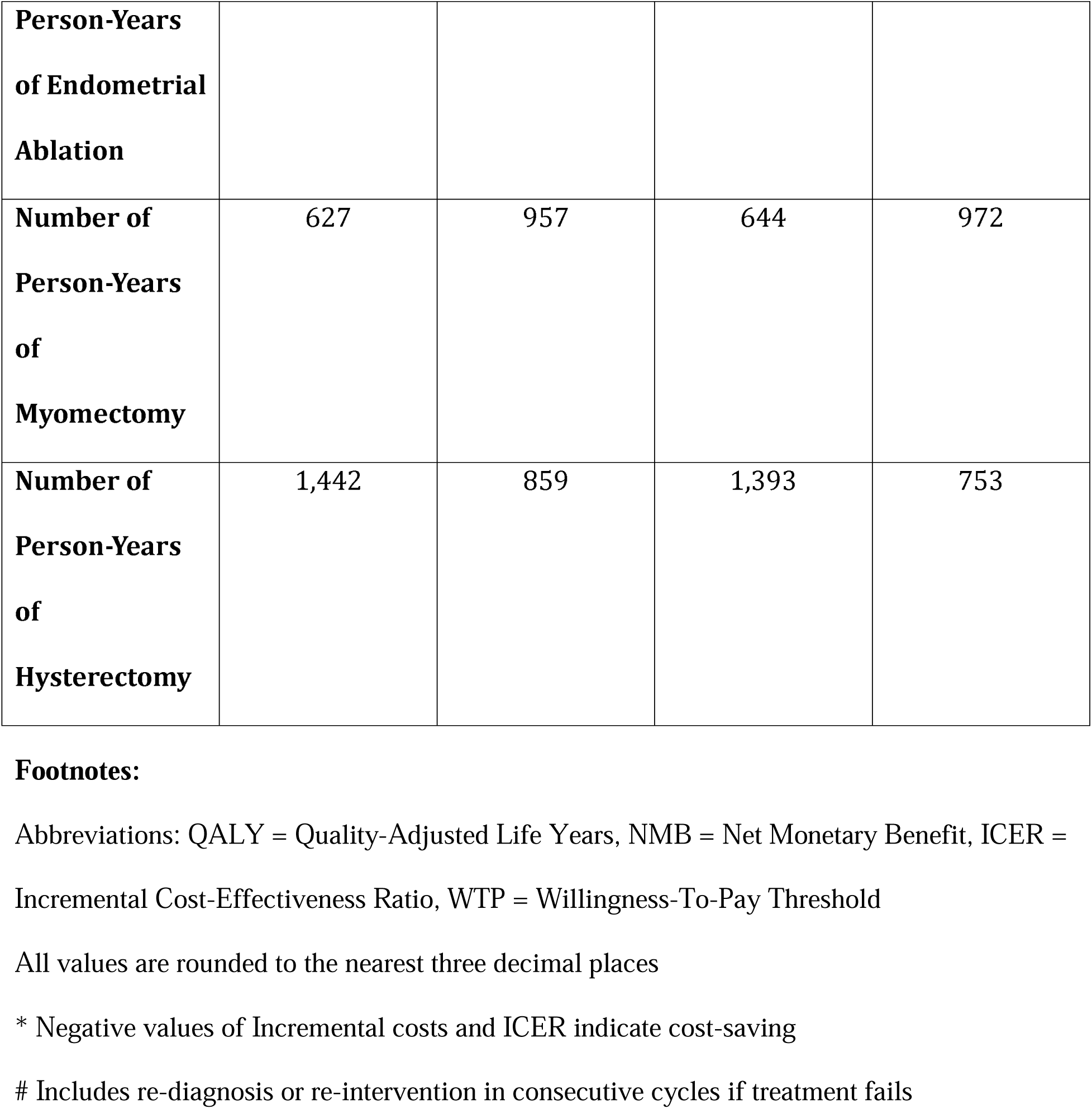
Base-Case Analysis Results of Cost-Effectiveness of Ultrasound Screening for Uterine Fibroids in the United States.

Univariate sensitivity analyses are visualized in Figure 2, a tornado diagram. These analyses determined the effect of variations in model variables (multiple univariate analyses) on the INMB with a willingness-to-pay threshold of $30,000/QALY. For both analyses (all women and Black women), the model was most sensitive to the probability of being diagnosed without having an ultrasound screening, followed by the cost of associated conditions in women with fibroids, the probability of symptomaticity, the cost of ultrasound screening, and the utilities in women treated successfully. For varying values of each of the model variables, the highest and lowest extremes of resultant INMBs in the univariate sensitivity analyses were $63,323.977 and $3,661.925 for all women, and $65,090.224 and $2,264.744 for Black women, respectively. The lowest values of INMB for all variables in both analyses were above zero, indicative of cost-effectiveness even in extremes of the ranges of model variables.

**Figure 2:**
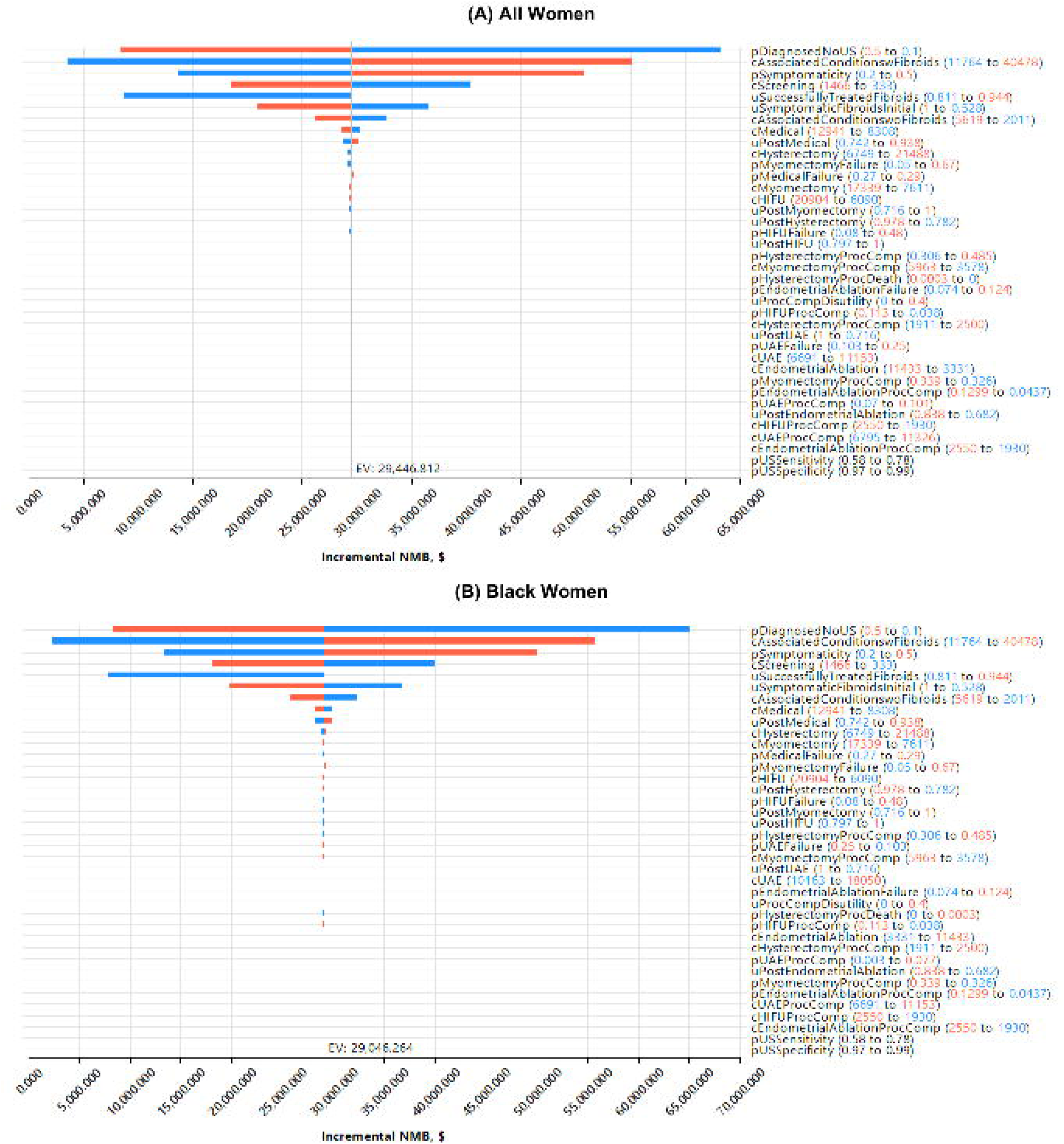
Tornado Diagram of Univariate Sensitivity Analyses of Cost-Effectiveness of Ultrasound Screening for Uterine Fibroids in the United States (A) All Women (B) Black Women. Tornado diagram shows multiple univariate analyses (effect of variations in one variable while keeping others constant) shown together in a comparative graph; Red side of each bar shows effect of higher values of each variable on incremental NMB and Blue side of each bar shows effect of lower values of each variable on incremental NMB. The model was most sensitive to the probability of being diagnosed without having an ultrasound screening, followed by the cost of associated conditions in women with fibroids, the probability of symptomaticity, utilities in women treated successfully, and utilities in women with symptomatic fibroids. **Abbreviations:** NMB = Net Monetary Benefit, HIFU = High-intensity Focused Ultrasound, UAE = Uterine Artery Embolization, ‘p’ = Probability of, ‘c’ = Costs of, ‘u’ = Utilities of, EV = Expected Value.

A univariate sensitivity analysis of the effect of starting age on screening showed that a starting age of 25 years is most cost-effective. As shown in Figure 3 and Appendix D, increasing the starting age of screening was associated with increasing incremental costs (decreasing cost-saving) and decreasing incremental utilities; overall leading to decreasing INMB of $27,306.429 (for starting at 25 years) to $1,194.465 (for a starting at 54 years). Similar trends were seen for ultrasound screening for Black women. These results indicated cost-effectiveness at all starting ages, with greater benefit at younger ages.

**Figure 3:**
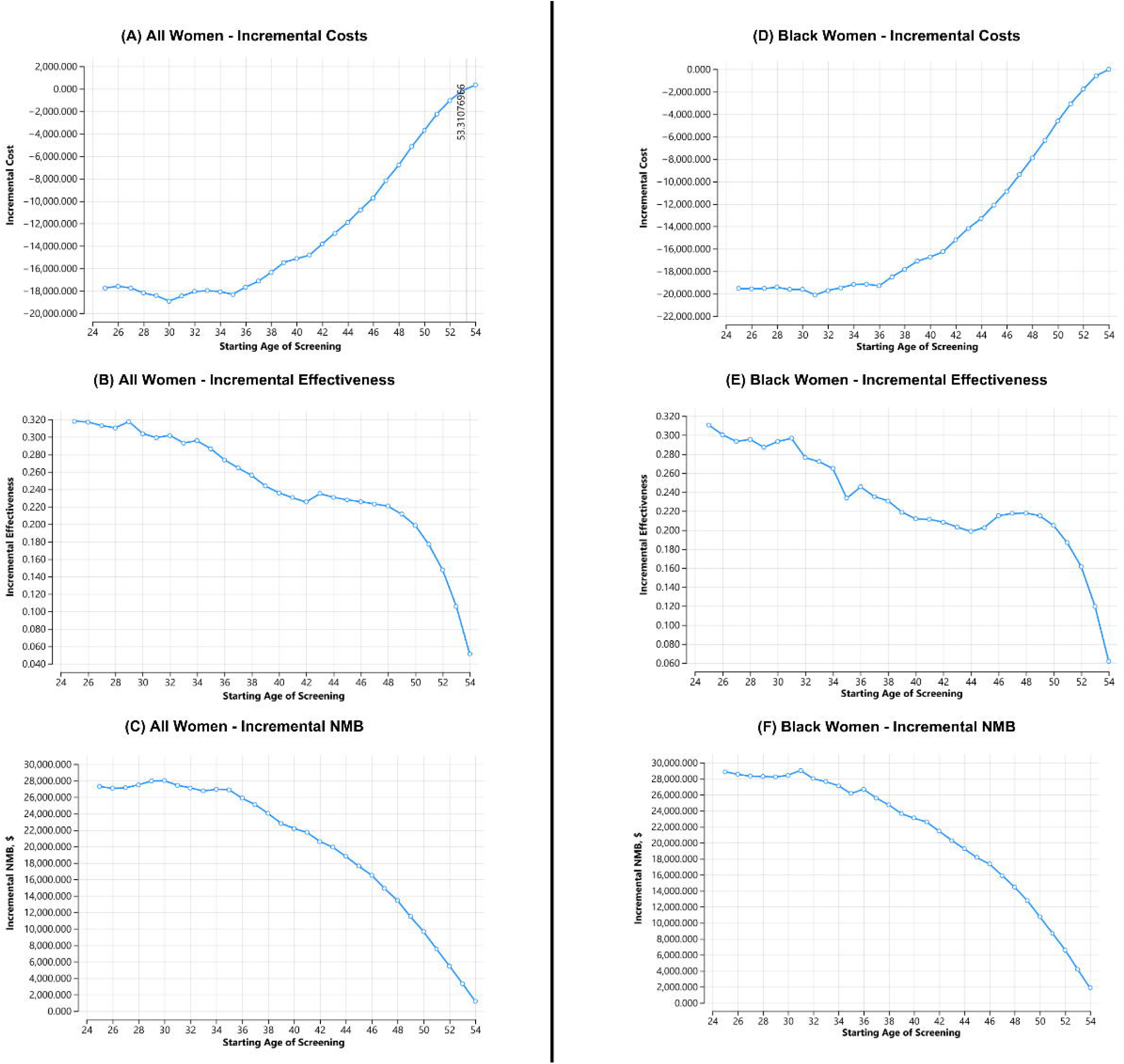
Effect of Starting Age of Screening on Cost-Effectiveness of Ultrasound Screening for Uterine Fibroids in the United States. (A, B, C) All Women: Incremental Costs, Incremental Effectiveness (Utilities), Incremental Net Monetary Benefit (D, E, F) Black Women: Incremental Costs, Incremental Effectiveness (Utilities), Incremental Net Monetary Benefit. Negative numbers on Y axis of incremental costs graphs indicate cost-saving (trends towards zero indicate lesser cost-saving with increasing starting age). Ultrasound screening for fibroids is more cost-saving when started at younger ages, with starting at 25 years being the most cost-effective. Screening is cost-effective at all ages of starting screening. **Abbreviations:** NMB = Net Monetary Benefit

Next, probabilistic sensitivity analyses were considered. Monte-Carlo simulations of 10,000 trials and 1,000 samples demonstrated that ultrasound screening was the preferred strategy for 92.3% of all women (Figure 4 A), considering both a negative incremental cost and a positive incremental effectiveness. In the 6.3% of the samples, a decrease in effectiveness was observed, which represents the cohort for which ultrasound screening is not a preferred strategy. Similarly, for Black women, ultrasound screening was found to be the preferred and dominant strategy for 91.2% of the cohort (Figure 4 B). For 6.9% of Black women, ultrasound screening had lower effectiveness.

**Figure 4:**
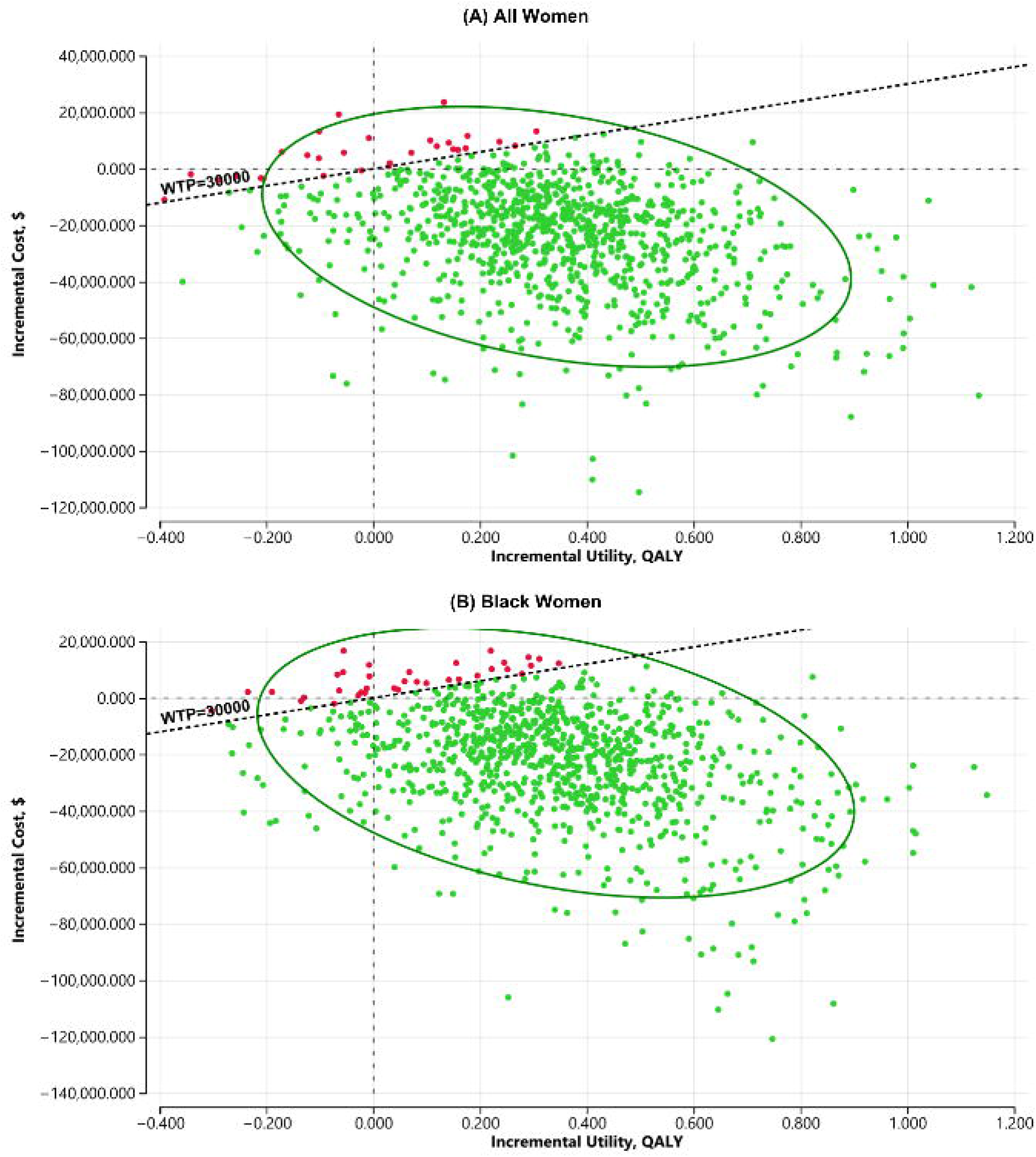
ICER Scatter Plot of Probabilistic Sensitivity Analyses of Cost-Effectiveness of Ultrasound Screening for Uterine Fibroids in the United States (A) All Women (B) Black Women. Negative numbers on Y axis indicate cost-saving, Green Dots represent the ICER (incremental cost effectiveness ratio) values for each of the 1000 samples, Ellipse represents 95% confidence interval, Bold dotted line represents willingness to pay threshold. Probabilistic sensitivity analyses demonstrated that ultrasound screening was the preferred strategy for 94.9% of all women and for 93.7% of Black women, compared to no screening. **Abbreviations:** WTP = Willingness-To-Pay Threshold, ICER = Incremental Cost Effectiveness Ratio

Through subsequent analysis for a population of 63.89 million women^65^ in the U.S. between the ages of 25 and 54 years, annual ultrasound screening for fibroids has the cumulative potential to save $1,169 billion and increase QALYs by 20.7 million compared to no ultrasound screening for fibroids (Refer to Appendix E for calculation). Similarly, for a subset population of 9.32 million Black women^65^ in the U.S. between the ages of 25 and 54 years, ultrasound screening could save $183 billion and increase QALYs by 3 million.

### Comment

#### Principal Findings

This Markov model-based cost-effectiveness evaluation demonstrated that annual ultrasound screening for fibroids was not only cost-effective but also cost-saving. Ultrasound screening led to earlier identification of women with symptomatic fibroids, with greater cost savings and QALY gains observed in Black women, who face a disproportionately higher burden of disease. Screening was cost-effective at all ages of starting screening. While most screening simulations demonstrated benefit, a minority of those suggested lower QALYs, possibly due to overdiagnosis and unnecessary interventions, underscoring the need for careful follow-up and shared decision-making.

#### Results in the Context of What is Known

Due to its high sensitivity and specificity,^23,24^ ultrasound screening is a highly accurate method for identifying women who are at elevated risk of uterine fibroids, thereby facilitating the opportunity for potential secondary prevention, early treatment or intervention.^20,21^ Although we found no prior cost-effectiveness analyses of routine fibroid screening, considerable evidence exists for the clinical burden of fibroids, longstanding diagnostic delays, and racial disparities. Studies have shown that many women experience delays in the diagnosis of uterine fibroids due to misattribution or dismissal of symptoms by healthcare providers or self-management of symptoms.^6^ These delays often result in more complex presentations, which typically require more extensive surgical management. In contrast, early diagnosis through ultrasound screening could significantly enhance the range of treatment options available to women.

Black women face a higher prevalence and symptom burden from uterine fibroids, making them an important sub-population to consider in the cost analysis of ultrasound screening.^2,8,9,30^ Evidence reveals that disparities in fibroid severity are exacerbated by disparate treatment options, and inequities in access to care.^1,10,11^ These considerations necessitated a separate analysis focusing on disparities in fibroid prevalence,^2,30^ while maintaining consistent parameters for symptomaticity and probability of diagnosis without a screening. The model demonstrated that ultrasound screening would be more cost-saving for Black women. This finding affirms the need for a disparities-conscious approach to healthcare analysis.

#### Clinical Implications

The impact of screening on clinical care is especially important if earlier detection may shape treatment choices and secondary prevention of disease progression. When detected earlier, women with fibroids may benefit from more prompt symptomatic management, including reducing fibroid size with gonadotropin-releasing hormone (GnRH) agonists and selective progestin receptor modulators, or providing symptomatic relief from heavy bleeding with levonorgestrel intrauterine devices (IUDs), combined oral contraceptive pills, non-steroidal anti-inflammatory drugs (NSAIDs), and tranexamic acid.^37^ Additionally, secondary prevention strategies, such as green tea or Vitamin D supplementation are low cost options that have been shown to reduce fibroid size or alleviate fibroid-associated symptoms.^67–70^ Minimally invasive, uterine-sparing techniques, including uterine artery embolization, myomectomy, endometrial ablation, magnetic resonance guided focused ultrasound surgery (MRgFUS), and radiofrequency ablation techniques show higher success rates and lower complication rates with treatment of smaller fibroids.^37,71,72^ However, the impact of these techniques on fertility and obstetric outcomes varies. For instance, myomectomies preserve fertility, and may enhance fertility depending on fibroid location,^37,71^ but may sometimes increase obstetric complications such as uterine rupture, preterm delivery, and cesarean section.^37^ Some reports indicate that uterine artery embolization is associated with both poorer fertility and obstetric outcomes, including lower postoperative pregnancy rates, increased miscarriage rates, cesarean section, and postpartum hemorrhage.^37,73,74^ The long-term impacts of fibroid radiofrequency ablation on fertility and obstetric outcomes remains to be determined,^74^ though studies are on-going.

Integrating ultrasound screening into routine care may aid early fibroid diagnosis and thus empower women with more effective and less invasive treatment options, balancing varying postoperative fertility and obstetric outcomes.

As with any population screening program, there is a possibility of false positives, overdiagnosis or unnecessary interventions that may not ultimately improve patient outcomes. However, by definition,^21^ screening should be viewed as an initial identification step among apparently healthy individuals that requires confirmatory evaluation before any treatment decisions are made. Similarly, it is crucial to address the lack of established guidelines for managing asymptomatic fibroids. Specifically, ACOG recommends expectant management when symptoms are not severe, with treatment decisions guided by shared decision-making.^25^ American Society for Reproductive Medicine (ASRM) recommends treatment for cavity-distorting or fertility-impacting fibroids, though these scenarios were not included in the model as stated in the methods.^75^ But, given that approximately 30% of fibroids may become symptomatic over time,^76^ a modified monitoring schedule and early referrals to fibroid specialists may be valuable. Moreover, widespread screening may reduce the normalization of symptoms by enabling patients to associate previously overlooked complaints with a confirmed diagnosis, facilitating earlier presentation and management.^14^ Collectively, a shared decision-making approach integrating patient preferences, symptoms, and fertility goals with provider expertise can lead to the selection of interventions with favorable risk-benefit profiles.

This study offers a robust foundation for developing guidelines to improve healthcare access, decrease diagnostic delays, and expand treatment options. However, given the model-based nature of the study, the findings of this study should be interpreted in context, as the model may not fully capture the nuances of real-world clinical practice. In addition, there are potential challenges to real-world implementation like variations in healthcare infrastructure, access to ultrasound technology, provider adherence, patient preferences, payer and reimbursement structures, and system-level administrative policy decisions. Given these and other unforeseen barriers, initial adoption could be pursued through pilot programs or phased implementation to assess feasibility and inform wider scale-up.

#### Research Implications

Owing to the uncertainty of available literature, the study adopted a conservative assumption that 30% ± 20% of unscreened symptomatic women with fibroids would be diagnosed.^33^ Costs and utilities related to infertility and indirect costs associated with the loss of productivity were also excluded. Despite these factors, the incremental net monetary benefit remained positive, suggesting applicability to real-world scenarios. Further research providing more refined data and reduced parameter uncertainty could help to strengthen future cost-effectiveness analyses of ultrasound screening for fibroids. Similarly, new minimally invasive techniques, such as radiofrequency ablation, are available clinically but are not included in the model due to insufficient data. The decision tree could also be adapted to include variables such as the effects of fibroids on infertility and pregnancy, and the impact of quicker referrals, alternative screening frequencies, medical and surgical management, and secondary prevention options.

#### Strengths and Limitations

The study has the inherent limitations of any cost-effectiveness analysis, including assumptions and uncertainties in model variables, such as ultrasonography operator-variance on accuracy, fibroid progression, symptomaticity, and patient preferences. For simplicity we assumed an annual screening which may not be the optimal screening interval for cost effectiveness and less frequent screening intervals could be examined. These limitations were mitigated through the inclusion of values from high-quality evidence, employment of appropriate mathematical techniques, and validation of all values and assumptions by clinical experts. While published data is ideal, this is an acceptable approach to filling in gaps in the literature for decision-analytic modeling.^77,78^

A major strength of this cost-effectiveness evaluation is the use of validated variables, payoff discounting, and probabilistic sensitivity analyses to report net monetary benefits for a large population-based screening trial. Additionally, the use of a sub-analysis for Black women, a group disproportionately impacted by fibroids, further strengthens the utility of this study. The study’s findings underscore the substantial cumulative benefits of screening strategies in gynecology and suggest avenues for improved cost-effectiveness in the U.S.

#### Conclusions

Based on this decision-analytic Markov model, ultrasound screening for uterine fibroids in women aged 25 to 54 years in the United States was cost-saving, even more so for Black women. These model-based findings highlight the potential value of developing the current clinical practices to include annual ultrasound screening for fibroids, which could aid in early diagnosis, potential secondary prevention, and timely intervention. The screening can have a significant positive cumulative impact on enhancing the quality of life of women with fibroids and decreasing costs for healthcare payers. Future research and pilot implementation studies are warranted to validate these results in real-world settings, assess feasibility, and determine their actual impact.

## Supporting information

Consolidated Health Economic Evaluation Reporting Standards 2022 (CHEERS 2022) Statement

Calculations and Derivations of Values of Model Parameters (Variables)

Decision Tree and Assumptions

Results of Alternative Analysis of Starting Age

Calculations of Cumulative Lifetime Economic and Health Outcomes

## Data Availability

The decision tree used for the cost-effectiveness evaluation has been shared in Figure 1 and Figure C.1. The model parameters used and all the derivations and other calculations of values have been shared in Table 1 and Appendix B.

## Glossary

- Willingness-to-pay (WTP) threshold: the maximum amount a healthcare system or society is considered willing to spend to gain one additional QALY, used as a benchmark to judge whether an intervention is cost-effective; often decided by national specialty organizations.
- Half-cycle correction: a correction that reduces bias by assuming events occur mid-cycle rather than only at the start or end, thereby better reflecting real-world scenarios
- Discounting of pay-offs: an adjustment that values future costs and health benefits slightly less than those occurring today, to reflect time preference and allow fair comparison across years.
- Incremental cost-effectiveness ratio (ICER): a measure that compares the difference in costs and health outcomes between two strategies, expressed as the additional cost required to gain one QALY.
- Incremental net monetary benefit (INMB): a measure that converts health benefits into monetary terms using a willingness-to-pay threshold (net monetary benefit), allowing the difference in total value between two strategies to be expressed as a single number, where positive values favor the intervention.

## Acknowledgments

The authors gratefully acknowledge Miriam Doyle-Baschat and the faculty and staff of the Reproductive Sciences Division in addition to administrative support from the Department of Gynecology and Obstetrics at Johns Hopkins and the assistance of the VISMED program. The authors acknowledge efforts and early discussions with Chelsea Henshaw and the BEAD Core at Johns Hopkins for initial conceptualization and discussions of methodology.

## Author Contributions: CRediT

**PM**: Methodology, Investigation, Formal analysis, Visualization, Writing – original draft; **LVR**: Methodology, Investigation, Formal analysis, Visualization, Writing – original draft; **AS**: Investigation, Visualization, Writing – original draft; **KS, KP, AF, MVV, HW, KW**: Validation; **BS**: Conceptualization, Methodology, Investigation, Formal analysis, Project administration, Resources, Supervision, Validation; **JHS**: Conceptualization, Methodology, Project administration, Resources, Supervision, Validation; **All authors**: Writing – review & editing.

## References

1. Huang D, Magaoay B, Rosen MP, Cedars MI. Presence of Fibroids on Transvaginal Ultrasonography in a Community-Based, Diverse Cohort of 996 Reproductive-Age Female Participants. JAMA Netw Open 2023; 6(5): e2312701. doi:10.1001/jamanetworkopen.2023.12701.

2. Stewart EA, Cookson CL, Gandolfo RA, Schulze-Rath R. Epidemiology of uterine fibroids: a systematic review. BJOG 2017; 124(10): 1501–12. doi:10.1111/1471-0528.14640.

3. Stewart EA, Laughlin-Tommaso SK, Catherino WH, Lalitkumar S, Gupta D, Vollenhoven B. Uterine fibroids. Nat Rev Dis Primers 2016; 2: 16043. doi:10.1038/nrdp.2016.43.

4. Shavell VI, Thakur M, Sawant A, et al. Adverse obstetric outcomes associated with sonographically identified large uterine fibroids. Fertil Steril 2012; 97(1): 107–10. doi:10.1016/j.fertnstert.2011.10.009.

5. Go VAA, Thomas MC, Singh B, et al. A systematic review of the psychosocial impact of fibroids before and after treatment. Am J Obstet Gynecol 2020; 223(5): 674,708.e8. doi:10.1016/j.ajog.2020.05.044.

6. Marsh EE, Al-Hendy A, Kappus D, Galitsky A, Stewart EA, Kerolous M. Burden, Prevalence, and Treatment of Uterine Fibroids: A Survey of U.S. Women. J Womens Health (Larchmt) 2018; 27(11): 1359–67. doi:10.1089/jwh.2018.7076.

7. Cheng LC, Li HY, Gong QQ, Huang CY, Zhang C, Yan JZ. Global, regional, and national burden of uterine fibroids in the last 30 years: Estimates from the 1990 to 2019 Global Burden of Disease Study. Front Med (Lausanne) 2022;9:1003605. Published 2022 Nov 7. doi:10.3389/fmed.2022.1003605

8. Wegienka G, Stewart EA, Nicholson WK, et al. Black Women Are More Likely Than White Women to Schedule a Uterine-Sparing Treatment for Leiomyomas. J Womens Health (Larchmt) 2021;30(3):355–66. doi:10.1089/jwh.2020.8634

9. Laughlin SK, Stewart EA. Uterine leiomyomas: individualizing the approach to a heterogeneous condition. Obstet Gynecol 2011 Feb;117(2 Pt 1):396–403. doi: 10.1097/AOG.0b013e31820780e3.

10. Bower JK, Schreiner PJ, Sternfeld B, Lewis CE. Black-White differences in hysterectomy prevalence: the CARDIA study. Am J Public Health 2009; 99(2): 300–7. doi:10.2105/AJPH.2008.133702.

11. Sengoba KS, Ghant MS, Okeigwe I, Mendoza G, Marsh EE. Racial/Ethnic Differences in Women’s Experiences with Symptomatic Uterine Fibroids: a Qualitative Assessment. J Racial Ethn Health Disparities 2017; 4(2): 178–83. doi:10.1007/s40615-016-0216-1.

12. Katon JG, Plowden TC, Marsh EE. Racial disparities in uterine fibroids and endometriosis: a systematic review and application of social, structural, and political context. Fertil Steril 2023; 119(3): 355–63. doi:10.1016/j.fertnstert.2023.01.022.

13. Harmon QE, Actkins KV, Baird DD. Fibroid Prevalence-Still So Much to Learn. JAMA Netw Open 2023; 6(5): e2312682. doi:10.1001/jamanetworkopen.2023.12682.

14. Ghant MS, Sengoba KS, Vogelzang R, Lawson AK, Marsh EE. An Altered Perception of Normal: Understanding Causes for Treatment Delay in Women with Symptomatic Uterine Fibroids. J Womens Health (Larchmt) 2016; 25(8): 846–52. doi:10.1089/jwh.2015.5531.

15. Hazimeh D, Coco A, Casubhoy I, Segars J, Singh B. The Annual Economic Burden of Uterine Fibroids in the United States (2010 Versus 2022): A Comparative Cost-Analysis. Reprod Sci 2024; 31(12): 3743–56. doi:10.1007/s43032-024-01727-0.

16. Wang A, Wang S, Owens CD, Vora JB, Diamond MP. Health Care Costs and Treatment Patterns Associated with Uterine Fibroids and Heavy Menstrual Bleeding: A Claims Analysis. J Womens Health (Larchmt) 2022;31(6):856–863. doi: 10.1089/jwh.2020.8983.

17. Kowada A. Cost-effectiveness and health impact of lung cancer screening with low-dose computed tomography for never smokers in Japan and the United States: a modelling study. BMC Pulm Med 2022;22(1):19. doi: 10.1186/s12890-021-01805-y

18. Goldie SJ, Gaffikin L, Goldhaber-Fiebert JD, et al. Cost-effectiveness of cervical-cancer screening in five developing countries. N Engl J Med 2005;353(20):2158–68. doi: 10.1056/NEJMsa044278.

19. Sharma M, John R, Afrin S, et al. Cost-Effectiveness of Population Screening Programs for Cardiovascular Diseases and Diabetes in Low- and Middle-Income Countries: A Systematic Review. Front Public Health 2022;10:820750. doi: 10.3389/fpubh.2022.820750

20. Raffle AE, Mackie A, Muir Gray JA. Screening : Evidence and Practice. 2nd ed. Oxford University Press; 2019.

21. Wilson JMG, Jungner G, World Health Organization. Principles and practice of screening for disease. 1968.

22. Marshall LM, Spiegelman D, Barbieri RL, et al. Variation in the incidence of uterine leiomyoma among premenopausal women by age and race. Obstet Gynecol 1997;90(6):967–73. doi:10.1016/S0029-7844(97)00534-6

23. Maheux-Lacroix S, Li F, Laberge PY, Abbott J. Imaging for Polyps and Leiomyomas in Women With Abnormal Uterine Bleeding: A Systematic Review. Obstet Gynecol. 2016 Dec;128(6):1425–36. doi: 10.1097/AOG.0000000000001776.

24. Dueholm M, Lundorf E, Hansen ES, Ledertoug S, Olesen F. Accuracy of magnetic resonance imaging and transvaginal ultrasonography in the diagnosis, mapping, and measurement of uterine myomas. Am J Obstet Gynecol 2002; 186(3): 409–15. doi:10.1067/mob.2002.121725.

25. Management of Symptomatic Uterine Leiomyomas: ACOG Practice Bulletin, Number 228. Obstet Gynecol 2021 Jun 1;137(6):e100–e115. doi: 10.1097/AOG.0000000000004401

26. De La Cruz MS, Buchanan EM. Uterine Fibroids: Diagnosis and Treatment. Am Fam Physician 2017 Jan 15;95(2):100–7

27. Zhang J, Go VA, Blanck JF, Singh B. A Systematic Review of Minimally Invasive Treatments for Uterine Fibroid-Related Bleeding. Reprod Sci 2022;29(10):2786–2809. doi: 10.1007/s43032-021-00722-z

28. Morris JM, Liang A, Fleckenstein K, Singh B, Segars J. A Systematic Review of Minimally Invasive Approaches to Uterine Fibroid Treatment for Improving Quality of Life and Fibroid-Associated Symptoms. Reprod Sci 2023;30(5):1495–1505. doi: 10.1007/s43032-022-01120-9

29. Wegienka G, Baird DD, Hertz-Picciotto I, et al. Self-reported heavy bleeding associated with uterine leiomyomata. Obstet Gynecol 2003 Mar;101(3):431–7. doi: 10.1016/s0029-7844(02)03121-6.

30. Laughlin SK, Schroeder JC, Baird DD. New directions in the epidemiology of uterine fibroids. Semin Reprod Med 2010;28(3):204–17. doi:10.1055/s-0030-1251477

31. Xu J, Murphy S, Kochanek K, Arias E. Mortality in the United States, 2021.; 2022. doi:10.15620/cdc:122516

32. Divakar H. Asymptomatic uterine fibroids. Best Pract Res Clin Obstet Gynaecol 2008;22(4):643–54. doi:10.1016/j.bpobgyn.2008.01.007

33. Eltoukhi HM, Modi MN, Weston M, Armstrong AY, Stewart EA. The health disparities of uterine fibroid tumors for African American women: a public health issue. Am J Obstet Gynecol 2014;210(3):194–9. doi:10.1016/j.ajog.2013.08.008

34. Lee DW, Gibson TB, Carls GS, Ozminkowski RJ, Wang S, Stewart EA. Uterine fibroid treatment patterns in a population of insured women. Fertil Steril 2009;91(2):566–74. doi:10.1016/J.FERTNSTERT.2007.12.004

35. Flynn M, Jamison M, Datta S, Myers E. Health care resource use for uterine fibroid tumors in the United States. Am J Obstet Gynecol 2006;195(4):955–64. doi:10.1016/J.AJOG.2006.02.020

36. Al-Hendy A, Lukes AS, Poindexter AN, et al. Treatment of Uterine Fibroid Symptoms with Relugolix Combination Therapy. N Engl J Med 2021;384(7):630–42. doi:10.1056/NEJMOA2008283

37. Segars JH, Parrott EC, Nagel JD, et al. Proceedings from the Third National Institutes of Health International Congress on Advances in Uterine Leiomyoma Research: comprehensive review, conference summary and future recommendations. Hum Reprod Update 2014;20(3):309–33. doi:10.1093/humupd/dmt058

38. Bansi-Matharu L, Gurol-Urganci I, Mahmood TA, Templeton A, van der Meulen JH, Cromwell DA. Rates of subsequent surgery following endometrial ablation among English women with menorrhagia: population-based cohort study. BJOG 2013;120(12):1500–7. doi: 10.1111/1471-0528.12319

39. Davis MR, Soliman AM, Castelli-Haley J, Snabes MC, Surrey ES. Reintervention Rates After Myomectomy, Endometrial Ablation, and Uterine Artery Embolization for Patients with Uterine Fibroids. J Womens Health (Larchmt) 2018 Oct;27(10):1204–1214. doi: 10.1089/jwh.2017.6752

40. Cain-Nielsen AH, Moriarty JP, Stewart EA, Borah BJ. Cost-effectiveness of uterine-preserving procedures for the treatment of uterine fibroid symptoms in the USA. J Comp Eff Res 2014;3(5):503–14. doi:10.2217/CER.14.32

41. Sharp HT. Overview of endometrial ablation - UpToDate. UpToDate. Published February 24, 2023. Accessed January 05, 2026. https://www.uptodate.com/contents/overview-of-endometrial-ablation#topicContent

42. Kong CY, Meng L, Omer ZB, et al. MRI-guided focused ultrasound surgery for uterine fibroid treatment: a cost-effectiveness analysis. AJR Am J Roentgenol 2014;203(2):361–71. doi:10.2214/AJR.13.11446

43. Minalt N, Canela CD, Marino S. Endometrial Ablation - StatPearls - NCBI Bookshelf. In: StatPearls [Internet]. StatPearls Publishing; 2023. Accessed January 05, 2026. https://www.ncbi.nlm.nih.gov/books/NBK459245/

44. Pelvic Ultrasound Cost - 2024 Healthcare Costs. Accessed January 05, 2026. https://health.costhelper.com/pelvic-ultrasounds.html

45. Cardozo ER, Clark AD, Banks NK, Henne MB, Stegmann BJ, Segars JH. The estimated annual cost of uterine leiomyomata in the United States. Am J Obstet Gynecol 2012;206(3):211.e1–211.e9. doi:10.1016/J.AJOG.2011.12.002

46. Bonafede MM, Pohlman SK, Miller JD, Thiel E, Troeger KA, Miller CE. Women with Newly Diagnosed Uterine Fibroids: Treatment Patterns and Cost Comparison for Select Treatment Options. Popul Health Manag 2018;21(S1):S13–S20. doi: 10.1089/pop.2017.0151

47. Miller JD, Lenhart GM, Bonafede MM, Basinski CM, Lukes AS, Troeger KA. Cost effectiveness of endometrial ablation with the NovaSure(®) system versus other global ablation modalities and hysterectomy for treatment of abnormal uterine bleeding: US commercial and Medicaid payer perspectives. Int J Womens Health 2015;7:59–73. doi:10.2147/IJWH.S75030

48. Nagendra D, Gutman SM, Koelper NC, Loza-Avalos SE, Sonalkar S, Schreiber CA, Harvie HS. Medical management of early pregnancy loss is cost-effective compared with office uterine aspiration. Am J Obstet Gynecol 2022;227(5):737.e1–737.e11. doi: 10.1016/j.ajog.2022.06.054

49. Rausch M, Lorch S, Chung K, Frederick M, Zhang J, Barnhart K. A cost-effectiveness analysis of surgical versus medical management of early pregnancy loss. Fertil Steril 2012;97(2):355–60. doi: 10.1016/j.fertnstert.2011.11.044

50. Waitzman NJ, Jalali A, Grosse SD. Preterm birth lifetime costs in the United States in 2016: An update. Semin Perinatol 2021;45(3). doi:10.1016/J.SEMPERI.2021.151390

51. Rae M, Cox C, Dingel H. Health costs associated with pregnancy, childbirth, and postpartum care - Peterson-KFF Health System Tracker. Published July 13, 2022. Accessed January 05, 2026. https://www.healthsystemtracker.org/brief/health-costs-associated-with-pregnancy-childbirth-and-postpartum-care/

52. Nissenson AR, Wade S, Goodnough T, Knight K, Dubois RW. Economic burden of anemia in an insured population. J Manag Care Pharm 2005;11(7):565–74. doi:10.18553/JMCP.2005.11.7.565

53. Soliman AM, Surrey E, Bonafede M, Nelson JK, Castelli-Haley J. Real-World Evaluation of Direct and Indirect Economic Burden Among Endometriosis Patients in the United States. Adv Ther 2018;35(3):408–423. doi: 10.1007/s12325-018-0667-3

54. Yeh JM, Hook EW, Goldie SJ. A refined estimate of the average lifetime cost of pelvic inflammatory disease. Sex Transm Dis 2003;30(5):369–78. doi:10.1097/00007435-200305000-00001

55. Fink JL. How Common Are Miscarriages? | Miscarriage Rates, Statistics & Risk. Healthgrades. Published December 15, 2020. Accessed January 05, 2026. https://www.healthgrades.com/right-care/pregnancy/how-common-is-miscarriage

56. Osterman MJK, Hamilton BE, Martin JA, Driscoll AK, Valenzuela CP. National Vital Statistics Reports Volume 72, Number 1 January 31, 2023.; 2021. https://www.cdc.gov/nchs/products/index.htm.

57. Fennessy FM, Kong CY, Tempany CM, Swan JS. Quality-of-life assessment of fibroid treatment options and outcomes. Radiology 2011;259(3):785–92. doi:10.1148/RADIOL.11100704/-/DC1

58. O’Sullivan AK, Thompson D, Chu P, Lee DW, Stewart EA, Weinstein MC. Cost-effectiveness of magnetic resonance guided focused ultrasound for the treatment of uterine fibroids. Int J Technol Assess Health Care 2009;25(1):14–25. doi:10.1017/S0266462309090035

59. Russell LB, Gold MR, Siegel JE, Daniels N, Weinstein MC. The Role of Cost-effectiveness Analysis in Health and Medicine. JAMA 1996;276(14):1172–7. doi:10.1001/JAMA.1996.03540140060028

60. Consumer Price Index Data from 1913 to 2025; Available at: https://www.usinflationcalculator.com/inflation/consumer-price-index-and-annual-percent-changes-from-1913-to-2008/. Accessed January 05, 2026.

61. Freytag D, Günther V, Maass N, Alkatout I. Uterine Fibroids and Infertility. Diagnostics (Basel) 2021; 11(8): 1455. doi: 10.3390/diagnostics11081455. doi:10.3390/diagnostics11081455.

62. TreeAge Software LLC. TreeAge Pro Healthcare [Computer Software]. 2025.

63. Naimark DMJ, Bott M, Krahn M. The half-cycle correction explained: two alternative pedagogical approaches. Med Decis Making 2008; 28(5): 706–12. doi:10.1177/0272989X08315241.

64. Appleby J, Devlin N, Parkin D. NICE’s cost effectiveness threshold. BMJ 2007; 335(7616): 358–9. doi:10.1136/bmj.39308.560069.BE.

65. US Census Bureau. Age and Sex Composition in the United States: 2020; Available at: https://www.census.gov/data/tables/2020/demo/age-and-sex/2020-age-sex-composition.html. Accessed January 05, 2026.

66. US Census Bureau. The Black Alone Population in the United States: 2019; Available at: https://www.census.gov/data/tables/2019/demo/race/ppl-ba19.html. Accessed January 05, 2026.

67. Grandi G, Del Savio MC, Melotti C, Feliciello L, Facchinetti F. Vitamin D and green tea extracts for the treatment of uterine fibroids in late reproductive life: a pilot, prospective, daily-diary based study. Gynecol Endocrinol 2022; 38(1): 63–7. doi:10.1080/09513590.2021.1991909.

68. Ciavattini A, Delli Carpini G, Serri M, et al. Hypovitaminosis D and "small burden" uterine fibroids: Opportunity for a vitamin D supplementation. Medicine (Baltimore) 2016;95(52):e5698. doi:10.1097/MD.0000000000005698

69. Vahdat M, Allahqoli L, Mirzaei H, et al. The effect of vitamin D on recurrence of uterine fibroids: A randomized, double-blind, placebo-controlled pilot study. Complement Ther Clin Pract 2022 Feb;46:101536. doi: 10.1016/j.ctcp.2022.101536. Epub 2022 Jan 24.

70. Biro R, Richter R, Ortiz M, Sehouli J, David M. Effects of epigallocatechin gallate-enriched green tea extract capsules in uterine myomas: results of an observational study. Arch Gynecol Obstet 2021;303(5):1235–43. doi:10.1007/s00404-020-05907-6

71. De Smit NS, de Lange ME, Boomsma MF, Huirnet JAF, Hehenkamp WJK. Current Treatment for Symptomatic Uterine Fibroids: Available Evidence and Therapeutic Dilemmas. Obstet Gynecol Surv 2026;81(1):15–16. doi: 10.1097/OGX.0000000000001484

72. Dominoni M, Gardella B, Gritti A, Pasquali MF, Spinillo A. Conservative Treatment of Uterine Myomas: A Network Meta-Analysis of Randomized Controlled Studies. J Minim Invasive Gynecol 2025;32(7):583–591.e1. doi: 10.1016/j.jmig.2024.12.012

73. Bucuri CE, Mihu D, Mălu an AM, Oprea AV, Roman MP, Ormindean CM, Nati ID, Suciu VE, Hăprean AE, Pavel A, Toma M, Ciortea R. Impact of Uterine Artery Embolization on Subsequent Fertility Outcomes: A Meta-Analysis of 20 Years of Clinical Evidence. J Clin Med 2025;14(20):7205. doi: 10.3390/jcm14207205

74. Cottrell CM, Stewart EA. Use of Uterine Artery Embolization for the Treatment of Uterine Fibroids: A Comparative Review of Major National Guidelines. J Minim Invasive Gynecol 2025;32(5):418–424. doi: 10.1016/j.jmig.2024.11.006

75. Practice Committee of the American Society for Reproductive Medicine. Removal of myomas in asymptomatic patients to improve fertility and/or reduce miscarriage rate: a guideline. Fertil Steril 2017 Sep;108(3):416–25. doi: 10.1016/j.fertnstert.2017.06.034

76. Giuliani E, As-Sanie S, Marsh EE. Epidemiology and management of uterine fibroids. Int J Gynaecol Obstet 2020; 149(1): 3–9. doi:10.1002/ijgo.13102.

77. Briggs AH, Weinstein MC, Fenwick EAL, et al. Model parameter estimation and uncertainty analysis: a report of the ISPOR-SMDM Modeling Good Research Practices Task Force Working Group-6. Med Decis Making 2012; 32(5): 722–32. doi:10.1177/0272989X12458348.

78. Weinstein MC, O’Brien B, Hornberger J, et al. Principles of good practice for decision analytic modeling in health-care evaluation: report of the ISPOR Task Force on Good Research Practices--Modeling Studies. Value Health 2003; 6(1): 9–17. doi:10.1046/j.1524-4733.2003.00234.x.

