## Supplementary material for "Cost-effectiveness of Ultrasound Screening for Uterine Fibroids in the United States": Calculations and Derivations of Values of Model Parameters (Variables)

### **Appendix B: Calculations and Derivations of Values of Model Parameters (Variables)**

This Appendix describes all variable values and the detailed methods of calculation of these values including base-case values, ranges, and distribution parameters, along with relevant assumptions.

#### Item 1: Time Horizon

The time horizon was chosen up to menopause because the risk of fibroids, its symptoms, and complications decrease drastically after menopause.^1^ A meta-analysis by Zhu et al. (2019) reported the mean age of menopause as 50.2 years with a standard deviation (SD) of 4.4 years.^2^ We targeted to account for the inclusion of 90% of the women. The Z score for the 90^th^ percentile is 1.282. Hence, the numerical upper limit of the time horizon was calculated as:

90^th^ percentile = Mean + Z_90_ x SD = Mean + 1.282 x SD = 50.2 + 1.282 x 4.4 = 50.2 + 5.6408 = 55.8 years ≈ 55 years.

#### Item 2: Prevalence of Uterine Fibroids in All Women at Various Ages in the US

Wegienka et al. (2003) reported values for women with fibroids in all women from ages 35 to 51 years, but did not report the percentages directly.^3^ Hence the following calculations were made:

**Table B.1. Calculation of Prevalence of Fibroids in All Women of Age Groups 35 to 51 Years From Data by Wegienka et al.^3^**

| **Age Group** | **Sample Size in Age Group (n)** | **Diagnosis of Focal Leiomyomata (n)** | **Calculation of Prevalence (Rounded Off)** |
| --- | --- | --- | --- |
| 35 – 40 years | 158 + 148 + 45 = 351 | 148 | 148 / 351 x 100 = 42.2% |
| 41 – 45 years | 107 + 196 + 38 = 341 | 196 | 196 / 341 x 100 = 57.5% |
| 46 – 51 years | 49 + 144 + 25 = 218 | 144 | 144 / 218 x 100 = 66.1% |

As values of other age groups were not available, assuming similar trends, they were calculated proportionally from data of the prevalence of fibroids in Black women by age.

Laughlin et al. (2010) reported values of prevalence (cumulative incidence) of fibroids in Black women in the form of a graph.^1^ Reading the graph gave the following approximate values:

**Table B.2. Prevalence (Cumulative Incidence) Of Fibroids in Black Women**

| **Age Group** | **Prevalence** |
| --- | --- |
| 25 – 30 years | 20% |
| 31 – 35 years | 35% |
| 36 – 40 years | 55% |
| 41 – 45 years | 70% |
| 46 – 50 years | 80% |
| >50 years | ≳ 80% |

For women above the age of 50 years, they reported that the prevalence could be above 80%. Hence, the prevalence of fibroids at ages 51 to 55 years was taken to be 80% as an assumption.

Using the trend in prevalence of fibroids in Black women above, the prevalence of fibroids in all women from ages 25 to 34 years and 55 years were calculated:

**Table B.3. Calculation of Prevalence of Fibroids in All Women of Ages 25 to 34 Years and 52 to 55 Years From Data by Wegienka et al.^3^ and Laughlin et al.^1^**

| **Age Group** | **Prevalence in All Women^3^** | **Prevalence in Black Women^1^** | **Calculation of Prevalence (Rounded Off)** |
| --- | --- | --- | --- |
| 25 – 29 years | - | 20% | 20 / 55 x 42.2 = 15.34% |
| 30 – 34 years | - | 35% | 35 / 55 x 42.2 = 26.85% |
| 35 – 40 years | 42.2% | 55% | - |
| 41 – 45 years | 57.5% | 70% | - |
| 46 – 51 years | 66.1% | 80% | - |
| 52 – 55 years | - | 80% | 80 / 80 x 66.1 = 66.1% |

**Prevalence of Fibroids by Age and Race**

**Table B.4. Prevalence of Fibroids in All Women as Compared to Black Women**

| **Age Group** | **Prevalence in All Women** | **Prevalence in Black Women** |
| --- | --- | --- |
| 25 – 29 years | 15.34% | 20% |
| 30 – 34 years | 26.85% | 35% |
| 35 – 40 years | 42.2% | 55% |
| 41 – 45 years | 57.5% | 70% |
| 46 – 51 years | 66.1% | 80% |
| 52 – 55 years | 66.1% | 80% |

#### Item 3: Background Death as per Age

There is a statistical possibility of death (due to various other reasons for a particular age group), which were accounted for in the decision tree, as ‘background death’. The Centers for Disease Control and Prevention - National Center for Health Statistics (CDC-NCHS) has published data of Mortality in the United States, 2021 in terms of deaths per 100,000 population.^4^ Hence, the probabilities were calculated as follows:

**Table B.5. Calculation of Probabilities of Background Death as per Age**

| **Age Group** | **Death rate per 100,000 population as per CDC^4^** | **Calculation of Probability in terms of percentage (Rounded Off)** |
| --- | --- | --- |
| 25 – 34 years | 180.8 | 180.8 / 100000 x 100 = 0.18% |
| 35 – 44 years | 287.9 | 287.9 / 100000 x 100 = 0.29% |
| 45 – 54 years | 531.0 | 531.0/ 100000 x 100 = 0.53% |

#### Item 4: Ultrasound Screening Parameters

Maheux-Lacroixet al., in their systematic review including 25 studies and 3213 women, reported a pooled sensitivity of 69% with a 95% confidence interval of 58-78% for ultrasound diagnosis of fibroids.^5^ By definition of sensitivity, this means that ultrasound screening, on an average, will detect fibroids in 69% of the women who have them.

Similarly, they reported a pooled specificity of 98% with a 95% confidence interval of 97-99% for ultrasound diagnosis of fibroids. This means, by definition, that ultrasound screening, on an average, will correctly show absence of fibroids in 98% of the women who do not have them.

#### Item 5: Symptomaticity

The data on the symptomaticity of fibroids has been highly variable and estimates have stated that the proportions of women with symptomatic fibroids are likely overestimated because of underreporting of asymptomatic fibroids. Numbers in the range of 20% to 50% have been reported by Divakar (2008)^6^, Eltoukhi et al. (2014)^7^, etc. Hence, we considered a conservative estimate of 30% for the base-case analysis. Sensitivity analyses performed with the range accounted for the uncertainty in the data.

#### Item 6: Treatment Choices at Various Ages

For a woman with symptomatic fibroids, various treatment options, like medical, surgical (uterine artery embolization, high-intensity focused ultrasound surgery, endometrial ablation, myomectomy, hysterectomy), or no treatment, have been available to choose from. Treatment choices cannot be taken uniformly across all ages because of multiple factors like the desire for future fertility, the time spent suffering with symptoms, etc. Hence, age-based treatment choices’ probabilities were decided to be applied. However, such values for the entire range of treatment options were not available. Hence, we calculated those by applying trends of rates of myomectomies (fertility-preserving) and hysterectomies (non-fertility-preserving) with assumptions of similarities of treatments. The method of the same is described as follows:

Amongst the various articles available, the data by Lee et al. (2009) was considered to be the most suitable as it contains most of the treatment options available.^8^ They reported the following treatment choice percentages:

**Table B.6. Percentages of Treatment Options as Reported by Lee et al.^8^**

| **Treatment Option** | **Percentage Reported** |
| --- | --- |
| All treatments | |
| Only surgical treatment | 16.8% |
| Both surgical and prescription drug treatment | 42.4% |
| Only prescription drug treatment | 22.4% |
| No surgical or prescription drug treatment | 18.4% |
| Total | 100% |
| Surgical treatments | |
| Hysterectomy only | 78.9% |
| Myomectomy only | 10.0% |
| Uterine artery embolization only | 3.0% |
| Endometrial ablation only | 4.1% |
| Multiple treatments | 4.0% |
| Total | 100% |

In the above data, the treatment of ‘both surgical and prescription drug treatment’ was equally split (42.4% / 2 = 21.2%) to distribute to surgical and medical treatment to derive the following percentages:

**Table B.7. Derivation of Probabilities of Broad Treatment Options for Age 45 Years**

| **Treatment Option** | **Percentage Reported** |
| --- | --- |
| Medical management | 22.4% + 21.2% = 43.6% |
| Surgical treatment | 16.8% + 21.2% = 38% |
| No surgical or prescription drug treatment | 18.4% |
| Total | 100% |

The percentage of surgical treatment options was split proportionate to the categorization reported by Lee et al., where the percentage of multiple treatments was assumedly allotted to HIFU (high-intensity focused ultrasound)/ MRgFUS (magnetic resonance-guided focused ultrasound) surgery due to lack of data.^8^ This assumption was valid as other narrow studies report similar probabilities of choosing HIFU.^9^ The calculations of surgical treatment options are:

**Table B.8. Derivation of Probabilities of Surgical Treatment Options for Age 45 Years**

| **Treatment Option** | **Percentage Reported** |
| --- | --- |
| UAE | 38 x 3% = 1.14% |
| HIFU/MRgFUS | 38 x 4% = 1.52% |
| Endometrial Ablation | 38 x 4.1% = 1.558% |
| Myomectomy | 38 x 10% = 3.8% |
| Hysterectomy | 38 x 78.9% = 29.982% |

As explained above, it would have been incorrect to apply these treatment choice probabilities to all ages on a generalized basis. The data above reported by Lee at al. had participants aged 25–54 years.^8^ The mean age of participants with significant leiomyomas was 42.5 years, and hence the above probabilities were considered applicable for the age of 45 years, summarized as follows:

**Table B.9. Probabilities of Treatment Choices for Age 45 Years**

| **Treatment Option** | **Percentage Reported** |
| --- | --- |
| No Treatment | 18.4% |
| Medical Management | 43.6% |
| UAE | 1.14% |
| HIFU/MRgFUS | 1.52% |
| Endometrial Ablation | 1.558% |
| Myomectomy | 3.8% |
| Hysterectomy | 29.982% |
| Total | 100% |

Whereas, Flynn et al. (2006) reported the rates of myomectomies and hysterectomies for various age groups in the form of graphs.^10^ Reading the graph gave the following values:

**Table B.10. Rates of Myomectomies and Hysterectomies as Reported by Flynn et al.^10^**

| **Age** | **Rate of myomectomies per 10000 women** | **Rate of hysterectomies per 10000 women** |
| --- | --- | --- |
| 25 years | 10 | 1 |
| 30 years | 30 | 40 |
| 35 years | 12 | 118 |
| 40 years | 7 | 125 |
| 45 years | 2 | 115 |
| 50 years | 1 | 72 |
| 55 years | 7 | 10 |

Further, we made assumptions of similarities in the context of fertility preservation. The probabilities (trend) of no treatment, medical management, HIFU/MRgFUS were assumed to be similar to myomectomy, as they preserve fertility. The probabilities (trend) of UAE and endometrial ablation were assumed to be similar to hysterectomy, as they impact future fertility. Thus, probabilities from Table B.9 were given a trend in accordance with rates of myomectomies and hysterectomies; where,

Percentage of Treatment X at Age Y = (Percentage of Treatment X at age 45 / Rate of Myomectomy or Hysterectomy at age 45) x Rate of Myomectomy or Hysterectomy at age Y.

Such calculations led to the following numbers:

**Table B.11. Trends for Probabilities of Treatment Choices by Age**

| **Treatment Choices** | **25 years** | **30 years** | **35 years** | **40 years** | **45 years** | **50 years** | **55 years** |
| --- | --- | --- | --- | --- | --- | --- | --- |
| No Intervention | 92.000% | 276.000% | 110.400% | 64.400% | 18.400% | 9.200% | 64.400% |
| Medical Management | 218.000% | 654.000% | 261.600% | 152.600% | 43.600% | 21.800% | 152.600% |
| UAE | 0.010% | 0.397% | 1.170% | 1.239% | 1.140% | 0.714% | 0.099% |
| HIFU/ MRgFUS | 7.600% | 22.800% | 9.120% | 5.320% | 1.520% | 0.760% | 5.320% |
| Endometrial Ablation | 0.014% | 0.542% | 1.599% | 1.693% | 1.558% | 0.975% | 0.135% |
| Myomectomy | 19.000% | 57.000% | 22.800% | 13.300% | 3.800% | 1.900% | 13.300% |
| Hysterectomy | 0.261% | 10.429% | 30.764% | 32.589% | 29.982% | 18.771% | 2.607% |
| Total | 336.884% | 1021.167% | 437.453% | 271.142% | 100.000% | 54.121% | 238.462% |

However, the total of treatment choice probabilities at each age went above 100%, which required adjustment of total to 100% and of individual values in proportion to that. This was done by dividing by the total for that age and multiplying by 100. These calculations led to the following numbers which were used in the decision tree (with no treatment taken as complementary in decision tree):

**Table B.12. Final Probabilities of Treatment Choices by Age**

| **Treatment Choices** | **25 years** | **30 years** | **35 years** | **40 years** | **45 years** | **50 years** | **55 years** |
| --- | --- | --- | --- | --- | --- | --- | --- |
| No Intervention | 27.31% | 27.03% | 25.24% | 23.75% | 18.40% | 17.00% | 27.01% |
| Medical Management | 64.71% | 64.04% | 59.80% | 56.28% | 43.60% | 40.28% | 63.99% |
| UAE | 0.00% | 0.04% | 0.27% | 0.46% | 1.14% | 1.32% | 0.04% |
| HIFU/ MRgFUS | 2.26% | 2.23% | 2.08% | 1.96% | 1.52% | 1.40% | 2.23% |
| Endometrial Ablation | 0.00% | 0.05% | 0.37% | 0.62% | 1.56% | 1.80% | 0.06% |
| Myomectomy | 5.64% | 5.58% | 5.21% | 4.91% | 3.80% | 3.51% | 5.58% |
| Hysterectomy | 0.08% | 1.02% | 7.03% | 12.02% | 29.98% | 34.68% | 1.09% |
| Total | 100.00% | 100.00% | 100.00% | 100.00% | 100.00% | 100.00% | 100.00% |

#### Item 7: Failure (including Recurrence) Probabilities

Every treatment option could lead to adequate relief/ recurrence-free successful treatment, failure (which included both treatment failure and recurrence) or procedural death. The failure (including recurrence) probabilities of most treatment options were taken from Segars et al. (2014) as follows.^11^ Medical management failure probability was assumed to be similar to the reported rate of one of pharmacological options.^12^ Endometrial ablation failure/ recurrence rate was taken as average of 12-month reintervention percentages from Bansi-Matharu et al and Davis et al^13,14^ The failure probability of no treatment was assumed to be 100% assuming no spontaneous resolution. The failure probability of hysterectomy treatment was assumed to be 0%, as it is the definitive cure of fibroids, except for rare cases which were assumedly excluded. Success probabilities were complementary to them.

**Table B.13. Failure Probabilities of Treatment Options**

| **Treatment Option** | **Failure (including recurrence rate)** |
| --- | --- |
| No treatment | 100.00%* |
| Medical Management | 28.00%* |
| UAE | 17.65% |
| HIFU/MRgFUS | 28.00% |
| Endometrial Ablation | 9.90% |
| Myomectomy | 36.00% |
| Hysterectomy | 0.00%* |

(*Assumptions as explained above)

Ranges for the same were taken from Segars et al.^11^, Levine, Everyday Health^12^, Bansi-Matharu et al^13^ and Davis et al^14^ as follows:

**Table B.14. Ranges of Failure Probabilities of Treatment Options**

| **Treatment Option** | **Failure (including recurrence rate)** |
| --- | --- |
| Medical Management | 27.00% - 29.00% |
| UAE | 10.30% - 25.00% |
| HIFU/MRgFUS | 8.00% - 48.00% |
| Endometrial Ablation | 7.40% - 12.40% |
| Myomectomy | 5.00% - 67.00% |

#### Item 8: Procedural Complication Probabilities

Certain percentage of individuals, undergoing a procedure, experience complications of the procedure and are associated with increased costs and decreased quality of life. Hence, we accounted for the complications. We assumed the probabilities of procedural complications of medical management to be zero. We also assumed that the risk of complications was same irrespective of fibroid size or number or other factors related to the variation in length of the same surgery or patient-related comorbidities, which we acknowledge as a limitation. The base-case values and ranges for the probabilities of procedural complications of UAE, HIFU, myomectomy, and hysterectomy were obtained directly from scientific literature (Kong et al. (2014)^15^, Cain-Neilsen et al. (2014)^9^). The base-case value for probabilities of procedural complications of endometrial ablation was derived as the sum of minor complication rates obtained from UpToDate (2023).^16^ This gave the following values and ranges:

**Table B.15. Complication Probabilities of Treatment Options**

| **Treatment** | **Probabilities Base-Case Value** | **Probabilities Range** |
| --- | --- | --- |
| Medical Management | 0.00 | N/A |
| UAE | 3.6^9^ | 0.30 – 7.70^9^ |
| HIFU/MRgFUS | 3.00^9^ | 0.80 – 8.30^9^ |
| Endometrial Ablation | 3.90^16^ | N/A |
| Myomectomy | 18.25^15^ | 17.60 – 18.90^15^ |
| Hysterectomy | 37.20^15^ | 20.6 – 38.50^15^ |

However, these probabilities did not account for the need of blood transfusions and hence, blood transfusion rates were added to the above numbers as shown as follows^11^:

**Table B.16.** **Modified Complication Probabilities of Treatment Options**

| **Treatment** | **Blood Transfusion Rate Added^11^** | **Modified Base-Case Value** | **Modified Range** |
| --- | --- | --- | --- |
| UAE | ~ 0% | 3.60% | 0.30% - 7.70% |
| HIFU/MRgFUS | ~ 3% | 6.00% | 3.80% - 11.30% |
| Endometrial Ablation | ~ 3% | 6.90% | N/A |
| Myomectomy | ~ 15% | 33.25% | 32.60% - 33.90% |
| Hysterectomy | ~ 10% | 47.20 % | 30.6% - 48.50% |

The ranges for endometrial ablation were not available and hence, were derived proportionally (assumed to vary similarly) to HIFU, with formulae.

**Table B.17.** **Derivation of Ranges of Complication Probabilities of Endometrial Ablation and Hysterectomy**

| **Treatment** | **Base-Case Value** | **Range** | **Lower limit of Range Calculation** | **Upper limit of Range Calculation** |
| --- | --- | --- | --- | --- |
| HIFU/MRgFUS | 6.00% | 3.80% - 11.30% | - | - |
| Endometrial Ablation | 6.90% | - | 6.90 x 3.80 / 6.00 = 4.37% | 6.90 x 11.30 / 6.00 = 12.99% |

#### Item 9: Procedural Death Probabilities

We used the following probabilities, similar to those used by Kong et al. (2014) in their cost-effectiveness analysis^15^:

**Table B.18.** **Procedural Death Probabilities Used By Kong et al.^15^**

| **Treatment Option** | **Procedural Death** |
| --- | --- |
| UAE | 0.00% |
| HIFU/MRgFUS | 0.00% |
| Myomectomy | 0.00% |
| Hysterectomy | 0.02% |

Procedural death probability for medical management was assumed to be zero, as it is not a procedure. Procedural death probability for endometrial ablation was assumed to be zero, due to lack of data. This was considered appropriate as Minalt et al. (2022) described it as minimally invasive with no data on procedure-related death.^17^

#### Item 10: Probability of Diagnosis versus No Diagnosis in Unscreened Symptomatic Women (Symptom Neglect/ Delay in Diagnosis)

According to Marsh et al. (2020), many women with fibroids had not initially associated their symptoms with fibroids and had been likely undiagnosed.^18^ Amongst women who had a delay in diagnosis of fibroids, 48% reported they had no previous knowledge of fibroids, and 55% said they were “just dealing” with their symptoms.^19^ Hence, we made the conservative assumption that 30% ± 20% of unscreened symptomatic women with fibroids will be diagnosed.

#### Item 11: Intervention Costs (Direct Costs)

Every intervention cost was inflated using the Medical Care Index of the Consumer Price Index (CPI-MCI)^20^ from the CPI value of annual average of the source year to the CPI-MCI value of annual average of 2025 (equal to 580.102) with the help of the U.S. Bureau of Labor Statistics’ CPI inflation Calculator.^21^

**Table B.19.** **Direct Intervention Cost Ranges (Inflation-Adjusted to June 2025)**

| **Intervention** | **Cost Ranges (US $) Non-Inflated** | **Source Reference(s) for Range** | **Year(s) of Source Reference(s)** | **CPI-MCI(s) of Source Year(s)^20^** | **Cost Ranges (US $) Inflation- Adjusted** |
| --- | --- | --- | --- | --- | --- |
| Ultrasound | 250-1100 | ^22^ | 2024 | 563.841 | 333 - 1,466 |
|  | 525 | ^22^ | 2024 | 563.841 | 700 |
| Medical Management | 5,563-8,665 | ^23^ | 2011 (2010 $) | 388.436 | 8,308 - 12,941 |
| UAE | 8,126-13,544 | ^15^ | 2014 | 435.292 | 10,829 - 18,050 |
|  | 6,805-12,863 | ^23^ | 2011 (2010 USD) | 388.436 | 10,163 - 19,210 |
| HIFU/MRgFUS | 5,955 – 15,686 | ^9^ | 2014 | 435.292 | 7,936 - 20,904 |
|  | 4,570-7,617 | ^15^ | 2014 | 435.292 | 6,090 - 10,151 |
| Endometrial Ablation | 4,943 | ^23^ | 2011 (2010 $) | 388.436 | - |
| Myomectomy | 7,018–13,011 | ^9^ | 2014 | 435.292 | 9,353 - 17,339 |
|  | 5,711-9,518 | ^15^ | 2014 | 435.292 | 7,611 - 12,685 |
| Hysterectomy | 11,982-16,549 | ^24^ | 2018 (2015 $) | 446.752 | 15,558 - 21,488 |
|  | 5,064-8,440 | ^15^ | 2014 | 435.292 | 6,749 - 11,248 |

The final collective cost range was created using the minimum and maximum adjusted costs for each intervention, and the average value of each range was considered for base-case analysis. The range for endometrial ablation was assumed to vary in proportion to HIFU/MRgFUS and calculated accordingly.

**Table B.20.** **Final Collective Cost Range and Base-Case Direct Intervention Costs (Inflation-Adjusted to June 2025)**

| **Intervention** | **Final Collective Cost Range (US $) Inflation-Adjusted** | **Cost (US $) for Base-Case Inflation Adjusted** |
| --- | --- | --- |
| Pelvic Ultrasound | 333 - 1,466 | 900 |
| Medical Management | 8,308 - 12,941 | 10,625 |
| UAE | 10,163 - 18,050 | 14,107 |
| HIFU/MRgFUS | 6,090 - 20,904 | 13,497 |
| Endometrial Ablation | 3,331 - 11,433 | 7,382 |
| Myomectomy | 7,611 - 17,339 | 12,475 |
| Hysterectomy | 6,749 - 21,488 | 14,119 |

#### Item 12: Procedural Complication Costs

The same CPI-MCI inflation, as above, was used to calculate inflation-adjusted cost ranges, and the base-case cost for each intervention was calculated as the average of the minimum and maximum adjusted costs. Since medical management is not a procedure, the procedural complications costs were assumed to be zero. Additionally, no data was found for the procedural complication cost for HIFU/MRgFUS, so it was assumed to be similar to Endometrial Ablation as they have similar complications.

**Table B.21. Procedural Complication Cost Ranges (Inflation-Adjusted to June 2025)**

| **Intervention - Procedural Complication Costs** | **Cost Ranges (US $) Non-Inflated** | **Source Reference(s) for Range** | **Year(s) of Source Reference(s)** | **CPI-MCI(s) of Source Year(s)^20^** | **Cost Ranges (US $) Inflation-Adjusted** |
| --- | --- | --- | --- | --- | --- |
| Medical Management | 0 | Assumption | - | - | - |
| UAE | 5,021 - 8,369 | ^15^ | 2014 | 435.292 | 6,691 - 11,153 |
| HIFU/MRgFUS | 1,486 – 1,964 | Assumption to be similar to Endometrial ablation | 2015 | 446.752 | 1,930 - 2,550 |
| Endometrial Ablation | 1,486 – 1,964 | ^25^ | 2015 | 446.752 | 1,930 - 2,550 |
| Myomectomy | 2,685 - 4,475 | ^15^ | 2014 | 435.292 | 3,578 – 5,963 |
| Hysterectomy | 1,472 – 1,926 | ^25^ | 2015 | 446.752 | 1,911 – 2,500 |

**Table B.22.** **Final Collective Cost Range and Base-Case Procedural Complication Costs (Inflation-Adjusted to June 2025)**

| **Intervention - Procedural Complication Costs** | **Final Collective Cost Range (US $) Inflation-Adjusted** | **Cost (US $) for Base-Case Inflation Adjusted** |
| --- | --- | --- |
| Medical Management | - | 0 |
| UAE | 6,691 - 11,153 | 8,922 |
| HIFU/MRgFUS | 1,930 - 2,550 | 2,332 |
| Endometrial Ablation | 1,930 - 2,550 | 2,332 |
| Myomectomy | 3,578 – 5,963 | 4,771 |
| Hysterectomy | 1,911 – 2,500 | 2,206 |

#### Item 13: Cost of Health States

Throughout the Markov Model, the target population could have transitioned between four health states: Alive with Fibroids (Symptomatic or Asymptomatic), Alive without Fibroids, Alive without Fibroids (Successfully Treated), or Death (Background/Procedural). Each health state had an associated cost for each additional year in that health state, assuming that there was no additional cost once transitioned into the Death health state.

**Table B.23. Associated Cost of Each Health State, Calculated by Costs of Associated Conditions of Fibroids**

| **Health State** | **Source Reference(s) for Range** | **Final Collective Cost Range (US $) Inflation-Adjusted** | **Cost (US $) for Base-Case Inflation Adjusted** |
| --- | --- | --- | --- |
| Alive with Fibroids (Symptomatic) | Calculated Below | 11,764 - 40,478 | 26,121 |
| Alive with Fibroids (Asymptomatic) | Calculated Below | 2,011 - 5,619 | 3,815 |
| Alive without Fibroids | Calculated Below | 2,011 - 5,619 | 3,815 |
| Alive without Fibroids (Successfully Treated) | Calculated Below | 2,011 - 5,619 | 3,815 |
| Death (Background/ Procedural) | Assumption | - | 0 |

Each additional year a woman lived with symptomatic fibroids incurred costs of conditions associated with fibroids. A woman with symptomatic fibroids has been shown to have a higher risk of spontaneous abortion, preterm delivery, cesarean delivery, anemia, endometriosis, inflammatory diseases, and menstruation disorders, as they were either results of the symptoms or vice versa. By taking the product of the risk of these conditions when symptomatic fibroids are present and their respective costs, this was the average cost that was accounted for when a woman continued to live with symptomatic fibroids. Since women could have these conditions even when fibroids were asymptomatic or absent, the product of the general risk of each condition and their respective costs were accounted for when a woman continued to live without symptomatic fibroids. However, the risk of having these conditions has been known to be much higher with symptomatic fibroids, leading to a greater associated cost the longer the woman’s symptomatic fibroids were present. While symptomatic fibroids have been shown to be associated with many more conditions, we have limited to the above conditions for the simplicity of the model and to maintain a conservative approach. We also assumed that the risk of associated conditions was same irrespective of fibroid size or number, which we acknowledge as a limitation.

The costs and events associated with pregnancy would be expected to occur only a few times in each woman's lifetime. Factoring the costs of pregnancy-related conditions annually would have wrongly overestimated the costs. To address this, we adjusted those costs using publicly available data on fertility rate. According to the United States Centers for Disease Control (CDC)^26^ and World Bank,^27^ the total fertility rate in the U.S. is approximately 1.6 children per woman. This implies that, on average, a woman gives birth 1.6 times during her reproductive lifespan. Accordingly, we have adjusted the originally considered annually-recurring the annual costs for pregnancy-related complications (cesarean section and preterm birth) by dividing by 30 (the time-horizon-span from ages 25 to 54) and multiplying them by 1.6. This correction yields a more accurate estimate of pregnancy-related costs.

The same CPI-MCI inflation as above was used to calculate inflation-adjusted cost ranges for each associated condition of fibroids:

**Table B.24.** **Cost Ranges for Associated Conditions of Fibroids (Inflation-Adjusted to June 2025)**

| **Associated Conditions of Fibroids** | **Cost Range (US $) Inflation-Adjusted** | **Source Reference** |
| --- | --- | --- |
| Spontaneous Abortion | 589 - 921 | ^28^ |
|  | 787 – 1,257 | ^29^ |
| Preterm Delivery | 54,922 – 80,691 | ^30^ |
| Cesarean Delivery | 28,720 | ^31^ |
| Anemia | 26,088 | ^32^ |
| Endometriosis | 8,490 – 57,327 | ^33^ |
| Inflammatory Diseases | 4,198 – 12,390 | ^34^ |
| Menstruation Disorders | 3,845 – 36,991 | ^35^ |

**Table B.25.** **Rates of Associated Conditions When Fibroids Are Present Versus NOT Present**

| **Associated Conditions of Fibroids** | **Rates of Condition when Fibroids are present** | **Source Reference** | **Rates of Condition when Fibroids are NOT present** | **Source Reference** |
| --- | --- | --- | --- | --- |
| Spontaneous Abortion | 12 x 1.6 = 19.2% | ^23,36^ | 12% | ^36^ |
| Preterm Delivery | 7.6 x 1.5 = 11.40% | ^23,37^ | 7.60% | ^37^ |
| Cesarean Delivery | 20.41 x 3.7 = 75.52% | ^23,37^ | 20.41% | ^37^ |
| Anemia | 22.02% | ^8^ | 3.34% | ^8^ |
| Endometriosis | 17.04% | ^8^ | 0.68% | ^8^ |
| Inflammatory Diseases | 22.45% | ^8^ | 5.40% | ^8^ |
| Menstruation Disorders | 55.48% | ^8^ | 8.22% | ^8^ |

**Table B.26.** **Minimum and Maximum Average Costs of Associated Conditions When Fibroids Are Present (Inflation-Adjusted to June 2025)**

| **Associated Conditions when Fibroids are Present** | **Cost due to Individual (Minimum Value) (US $) Inflation-Adjusted** | **Cost due to Individual (Maximum Value) (US $) Inflation-Adjusted** |
| --- | --- | --- |
| Spontaneous Abortion* | 0.192 x 589 x 1.6 / 30 = 6.03 | 0.192 x 1,257 x 1.6 / 30 = 12.87 |
| Preterm Delivery* | 0.114 x 54,922 x 1.6 / 30 = 333.93 | 0.114 x 80,691 x 1.6 / 30 = 490.60 |
| Cesarean Delivery* | 0.7552 x 28,720 x 1.6 / 30 = 1,156.77 | 0.7552 x 28,720 x 1.6 / 30 = 1,156.77 |
| Anemia | 0.2202 x 26,088 = 5,744.58 | 0.2202 x 26,088 = 5,744.58 |
| Endometriosis | 0.1704 x 8,490 = 1,446.70 | 0.1704 x 57,327 = 9,768.52 |
| Inflammatory Diseases | 0.2245 x 4,198 = 942.45 | 0.2245 x 12,390 = 2,781.56 |
| Menstruation Disorders | 0.5548 x 3,845 = 2,133.21 | 0.5548 x 36,991 = 20,522.61 |
| Total | 11,764 | 40,478 |

*The pregnancy-related complications (preterm delivery and cesarean delivery) costs are adjusted by dividing by 30 (the time horizon span from ages 25 to 54) and multiplying them by 1.6, to avoid considering them as annually-recurring costs.

The base-case cost of associated conditions when fibroids are present is the average of the minimum and maximum costs: (11,764 + 40,478)/2 = $26,121

**Table B.27.** **Minimum and Maximum Average Costs of Associated Conditions When Fibroids Are NOT Present (Inflation-Adjusted to June 2025)**

| **Associated Conditions when Fibroids are NOT Present** | **Cost due to Individual (Minimum Value) (US $) Inflation-Adjusted** | **Cost due to Individual (Maximum Value) (US $) Inflation-Adjusted** |
| --- | --- | --- |
| Spontaneous Abortion* | 0.12 x 589 x 1.6 / 30 = 3.77 | 0.12 x 1,257 x 1.6 / 30 = 8.04 |
| Preterm Delivery* | 0.076 x 54,922 x 1.6 / 30 = 222.62 | 0.076 x 80,691 x 1.6 / 30 = 327.07 |
| Cesarean Delivery* | 0.2041 x 28,720 x 1.6 / 30 = 312.63 | 0.2041 x 28,720 x 1.6 / 30 = 312.63 |
| Anemia | 0.0334 x 26,088 = 871.34 | 0.0334 x 26,088 = 871.34 |
| Endometriosis | 0.0068 x 8,490 = 57.73 | 0.0068 x 57,327 = 389.82 |
| Inflammatory Diseases | 0.054 x 4,198 = 226.69 | 0.054 x 12,390 = 669.06 |
| Menstruation Disorders | 0.0822 x 3,845 = 316.06 | 0.0822 x 36,991 = 3,040.66 |
| Total | 2,011 | 5,619 |

*The pregnancy-related complications (preterm delivery and cesarean delivery) costs are adjusted by dividing by 30 (the time horizon span from ages 25 to 54) and multiplying them by 1.6, to avoid considering them as annually-recurring costs.

The base-case cost of associated conditions when fibroids are NOT present is the average of the minimum and maximum costs: (2,011 + 5,619)/2 = $3,815

#### Item 14: Utilities for Each Health State

Utilities are measured by Quality-Adjusted Life Years (QALYs), which is a standardized measure of disease burden. It combines length of life and quality of life into a single number between 0 and 1, where 1 is a perfect health state and 0 is a dead health state. The utility scores for each health state were taken as follows:

**Table B.28. Utilities for Each Health State Measured in QALYs**

| **Health State** | **Utility (QALY) for Base-Case Analysis** | **Source Reference for Base-Case** | **Utility (QALY) Range** | **Source Reference for Range** |
| --- | --- | --- | --- | --- |
| Alive with Fibroids (Symptomatic) | 0.743 | ^15,38,39^ (Average) | 0.528 - 1 | ^15,38,39^ |
| Alive with Fibroids (Asymptomatic) | 1.000 | Assumption | - | - |
| Alive without Fibroids | 1.000 | Assumption | - | - |
| Alive without Fibroids (Successfully Treated) | 0.944 | ^15,39^ | 0.811 - 0.944 | ^15,39^ |
| Death (Background/Procedural) | 0.000 | ^15^ | - | - |

#### Item 15: Utilities Following Successful Treatment without Procedural Complications

A woman with symptomatic fibroids may have improved their utility weight by receiving one of the many treatment options for fibroids. If the intervention was successful (without complications), then the woman’s utility was increased to the utilities listed below, depending on the treatment choice. However, this utility would have only been accounted for within that cycle/year, since the woman would have continued again at the “Successfully Treated” Health State.

**Table B.29.** **Utilities Following Successful Treatment (Only for That Year/Cycle)**

| **Treatment** | **Utility (QALY) for Base-Case Analysis** | **Source Reference for Base-Case** | **Utility (QALY) Range** | **Source Reference for Range** |
| --- | --- | --- | --- | --- |
| Medical Management Success | 0.840 | ^25^ | 0.742 – 0.938 | ^25^ |
| UAE Success | 0.920 | ^9^ | 0.716 - 1.000 | ^9^ |
| HIFU/MRgFUS Success | 0.925 | ^9^ | 0.797 - 1.000 | ^9^ |
| Endometrial Ablation Success | 0.760 | ^25^ | 0.682 – 0.838 | ^25^ |
| Myomectomy Success | 0.925 | ^9^ | 0.716 - 1.000 | ^9^ |
| Hysterectomy Success | 0.880 | ^25^ | 0.782 – 0.978 | ^25^ |

#### Item 16: Utilities Following Treatment with Procedural Complications

If the intervention a woman receives had complications, a 20% decrease in utility score was accounted for, compared to utilities of success with no complications.

#### Item 17: Disutility per Cycle for Living with Symptomatic Fibroids- Logistic Decay

If a woman had symptomatic fibroids but received no treatment, there was an added disutility per cycle to account for the worsening of symptoms over time. The longer a woman went undiagnosed or received no treatment, the lower her utility score was accounted to be. This decrease in utility was modeled by a logistic decay function, determined by:

*Value in the n^th^ cycle = L + (U – L)[(1/1+e^k(x-15)^)]*, where:

U = Upper bound, base-case utility for symptomatic fibroids/ value in the first cycle

L = Lower bound/asymptote, based on published evidence and clinical expert validation = 0.528^15,38,39^

k = 0.5 (steepness parameter chosen to fit the above-mentioned values)

#### Item 18: Probabilistic Sensitivity Analysis- Mean and Standard Deviation

Probabilistic sensitivity analysis (PSA) is an economic modeling technique that helps to quantify the level of confidence in a model, allocating uncertainty in the model’s output to different sources of uncertainty in its inputs. PSA was run by assigning each input (probability, cost, utility, etc.) a probability distribution. The distribution was either assigned as uniform, triangular, beta (mainly for utilities), or gamma (mainly for costs). For beta and gamma distributions, different parameters specific to the type of distribution were calculated automatically by TreeAge Pro with the inputs of mean and standard deviation (SD) of each input’s range. The base-case value was taken as the mean for every input, and the standard deviation was calculated using the following equation (with an assumption that ranges reported in literature with 95% confidence intervals):

Upper Limit = Mean + 1.96 x SD; and; Lower Limit = Mean - 1.96 x SD

Hence, $SD=\frac{Upper Limit - Lower Limit}{1.96 \times2}$ where 1.96 is the Z-score for 95% confidence interval.

**Table B.30.** **Calculations of SD for Each Variable That Required a Beta or Gamma Distribution**

| **Variable** | **Range (US $ for Costs/ QALYs for Utilities)** | **Calculation** | **Standard Deviation** |
| --- | --- | --- | --- |
| Cost of a Pelvic Ultrasound | 333 - 1,466 | (1,466-333)/(1.96x2) | 289 |
| Cost of Medical Management | 8,308 - 12,941 | (12,941-8,308)/(1.96x2) | 1,182 |
| Cost of UAE | 10,163 - 18,050 | (18,050-10,163)/(1.96x2) | 2,012 |
| Cost of HIFU/MRgFUS | 6,090 - 20,904 | (20,904-6,090)/(1.96x2) | 3,779 |
| Cost of Endometrial Ablation | 3,331 - 11,433 | (11,433-3,331)/(1.96x2) | 2,067 |
| Cost of Myomectomy | 7,611 - 17,339 | (17,339-7,611)/(1.96x2) | 2,482 |
| Cost of Hysterectomy | 6,749 - 21,488 | (21,488-6,749)/(1.96x2) | 3,760 |
| Cost of UAE Procedural Complications | 6,691 - 11,153 | (11,153-6,691)/(1.96x2) | 1,138 |
| Cost of HIFU/MRgFUS Procedural Complications | 1,930 - 2,550 | (2,550-1,930)/(1.96x2) | 158 |
| Cost of Endometrial Ablation Procedural Complications | 1,930 - 2,550 | (2,550-1,930)/(1.96x2) | 158 |
| Cost of Myomectomy Procedural Complications | 3,578 – 5,963 | (5,963-3,578)/(1.96x2) | 608 |
| Cost of Hysterectomy Procedural Complications | 1,911 – 2,500 | (2,500-1,911)/(1.96x2) | 150 |
| Cost of Living with Fibroids (Symptomatic) | 11,764 - 40,478 | (40,478-11,764)/(1.96x2) | 7,325 |
| Cost of Living with Fibroids (Asymptomatic)/Living without Fibroids/Living without Fibroids (Successfully Treated) | 2,011 - 5,619 | (5,619-2,011)/(1.96x2) | 920 |
| Utility of Living with Fibroids (Symptomatic) | 0.528 - 1.000 | (1.000-0.528)/(1.96x2) | 0.120 |
| Utility of Living without Fibroids (Successfully Treated) | 0.811 - 0.944 | (0.944-0.811)/(1.96x2) | 0.034 |
| Utility of Medical Management Success | 0.742 - 0.938 | (0.938-0.742)/(1.96x2) | 0.050 |
| Utility of UAE Success | 0.716 - 1.000 | (1.000-0.716)/(1.96x2) | 0.072 |
| Utility of HIFU/MRgFUS Success | 0.797 - 1.000 | (1.000-0.797)/(1.96x2) | 0.052 |
| Utility of Endometrial Ablation Success | 0.682 - 0.838 | (0.838-0.682)/(1.96x2) | 0.040 |
| Utility of Myomectomy Success | 0.716 - 1.000 | (1.000-0.716)/(1.96x2) | 0.072 |
| Utility of Hysterectomy Success | 0.782 - 0.978 | (0.978-0.782)/(1.96x2) | 0.050 |

#### Item 19: Method of Calculating Net Monetary Benefit

A base-case analysis in a Markov model, involves calculation of the expected costs, QALYs, and health outcomes per woman by aggregating transition probabilities through the decision tree.

Incremental Cost-Effectiveness Ratio (ICER) is calculated as:

ICER = Difference in Costs / Difference in QALYs

Similarly, net monetary benefit (NMB) is then calculated for each strategy is calculated as:

NMB = (QALYs x Willingness-to-Pay Threshold) – Cost

This is applied for each of the strategies separately.

The incremental net monetary benefit (INMB) is derived, based on difference in QALYs and costs between two strategies.

INMB = (Difference in QALYs x Willingness-to-Pay Threshold) – Difference in Costs

All these values are directly provided by the analytical software - TreeAge Pro. Being standard calculations in cost-effectiveness analyses, these formulae are not added to the main text; but are mentioned here for clarity to readers.

#### References to Appendix B

1.	Laughlin SK, Schroeder JC, Baird DD. New directions in the epidemiology of uterine fibroids. *Semin Reprod Med*. 2010;28(3):204-217. doi:10.1055/s-0030-1251477

2.	Zhu D, Chung HF, Dobson AJ, et al. Age at natural menopause and risk of incident cardiovascular disease: a pooled analysis of individual patient data. *Lancet Public Health*. 2019;4(11):e553-e564. doi:10.1016/S2468-2667(19)30155-0

3.	Wegienka G, Baird DD, Hertz-Picciotto I, et al. Self-reported heavy bleeding associated with uterine leiomyomata. *Obstetrics and Gynecology*. 2003;101(3):431-437. doi:10.1016/S0029-7844(02)03121-6

4.	Xu J, Murphy S, Kochanek K, Arias E. *Mortality in the United States, 2021*.; 2022. doi:10.15620/cdc:122516

5.	Maheux-Lacroix S, Li F, Laberge PY, Abbott J. Imaging for Polyps and Leiomyomas in Women With Abnormal Uterine Bleeding: A Systematic Review. *Obstet Gynecol*. 2016 Dec;128(6):1425–36. doi: 10.1097/AOG.0000000000001776.

6.	Divakar H. Asymptomatic uterine fibroids. *Best Pract Res Clin Obstet Gynaecol*. 2008;22(4):643-654. doi:10.1016/j.bpobgyn.2008.01.007

7.	Eltoukhi HM, Modi MN, Weston M, Armstrong AY, Stewart EA. The health disparities of uterine fibroid tumors for African American women: a public health issue. *Am J Obstet Gynecol*. 2014;210(3):194-199. doi:10.1016/j.ajog.2013.08.008

8.	Lee DW, Gibson TB, Carls GS, Ozminkowski RJ, Wang S, Stewart EA. Uterine fibroid treatment patterns in a population of insured women. *Fertil Steril*. 2009;91(2):566-574. doi:10.1016/J.FERTNSTERT.2007.12.004

9.	Cain-Nielsen AH, Moriarty JP, Stewart EA, Borah BJ. Cost-effectiveness of uterine-preserving procedures for the treatment of uterine fibroid symptoms in the USA. *J Comp Eff Res*. 2014;3(5):503-514. doi:10.2217/CER.14.32

10.	Flynn M, Jamison M, Datta S, Myers E. Health care resource use for uterine fibroid tumors in the United States. *Am J Obstet Gynecol*. 2006;195(4):955-964. doi:10.1016/J.AJOG.2006.02.020

11.	Segars JH, Parrott EC, Nagel JD, et al. Proceedings from the Third National Institutes of Health International Congress on Advances in Uterine Leiomyoma Research: comprehensive review, conference summary and future recommendations. *Hum Reprod Update*. 2014;20(3):309-333. doi:10.1093/humupd/dmt058

12.	Al-Hendy A, Lukes AS, Poindexter AN, et al. Treatment of Uterine Fibroid Symptoms with Relugolix Combination Therapy. *New England Journal of Medicine*. 2021;384(7):630-642. doi:10.1056/NEJMOA2008283

13.	Bansi-Matharu L, Gurol-Urganci I, Mahmood TA, Templeton A, van der Meulen JH, Cromwell DA. Rates of subsequent surgery following endometrial ablation among English women with menorrhagia: population-based cohort study. *BJOG* 2013;120(12):1500-7. doi: 10.1111/1471-0528.12319

14.	Davis MR, Soliman AM, Castelli-Haley J, Snabes MC, Surrey ES. Reintervention Rates After Myomectomy, Endometrial Ablation, and Uterine Artery Embolization for Patients with Uterine Fibroids. *J Womens Health (Larchmt)* 2018;27(10):1204-1214. doi: 10.1089/jwh.2017.6752

15.	Kong CY, Meng L, Omer ZB, et al. MRI-guided focused ultrasound surgery for uterine fibroid treatment: a cost-effectiveness analysis. *AJR Am J Roentgenol*. 2014;203(2):361-371. doi:10.2214/AJR.13.11446

16.	Sharp HT. Overview of endometrial ablation - UpToDate. UpToDate. Published February 24, 2023. Accessed October 23, 2023. https://www.uptodate.com/contents/overview-of-endometrial-ablation#topicContent

17.	Minalt N, Canela CD, Marino S. Endometrial Ablation - StatPearls - NCBI Bookshelf. In: *StatPearls [Internet].* StatPearls Publishing; 2023. Accessed October 23, 2023. https://www.ncbi.nlm.nih.gov/books/NBK459245/

18.	Understanding racial disparities for women with uterine fibroids. Accessed July 25, 2023. https://ihpi.umich.edu/news/understanding-racial-disparities-women-uterine-fibroids

19.	Ghant MS, Sengoba KS, Vogelzang R, Lawson AK, Marsh EE. An Altered Perception of Normal: Understanding Causes for Treatment Delay in Women with Symptomatic Uterine Fibroids. *J Womens Health (Larchmt)*. 2016;25(8):846-852. doi:10.1089/JWH.2015.5531

20.	Consumer Price Index Data from 1913 to 2023. Accessed October 23, 2023. https://www.usinflationcalculator.com/inflation/consumer-price-index-and-annual-percent-changes-from-1913-to-2008/

21.	CPI Inflation Calculator. Accessed October 23, 2023. https://www.bls.gov/data/inflation_calculator.htm

22.	Pelvic Ultrasound Cost - 2024 Healthcare Costs. Accessed June 05, 2025. https://health.costhelper.com/pelvic-ultrasounds.html

23.	Cardozo ER, Clark AD, Banks NK, Henne MB, Stegmann BJ, Segars JH. The estimated annual cost of uterine leiomyomata in the United States. *Am J Obstet Gynecol*. 2012;206(3):211.e1-211.e9. doi:10.1016/J.AJOG.2011.12.002

24.	Bonafede MM, Pohlman SK, Miller JD, Thiel E, Troeger KA, Miller CE. Women with Newly Diagnosed Uterine Fibroids: Treatment Patterns and Cost Comparison for Select Treatment Options. *Popul Health Manag* 2018;21(S1):S13-S20. doi: 10.1089/pop.2017.0151.

25.	Miller JD, Lenhart GM, Bonafede MM, Basinski CM, Lukes AS, Troeger KA. Cost effectiveness of endometrial ablation with the NovaSure(®) system versus other global ablation modalities and hysterectomy for treatment of abnormal uterine bleeding: US commercial and Medicaid payer perspectives. *Int J Womens Health*. 2015;7:59-73. doi:10.2147/IJWH.S75030

26.	Hamilton BE, Martin JA, Osterman MJK. Births: provisional data for 2023. Vital Statistics Rapid Release; no 35. April 2024. doi: 10.15620/cdc/151797

27.	World Development Indicators – Total Fertility Rate (births per woman), World Development Indicators, World Bank Group Archives, Washington, D.C., United States. Available at: https://data.worldbank.org/indicator/SP.DYN.TFRT.IN

28.	Nagendra D, Gutman SM, Koelper NC, Loza-Avalos SE, Sonalkar S, Schreiber CA, Harvie HS. Medical management of early pregnancy loss is cost-effective compared with office uterine aspiration. *Am J Obstet Gynecol* 2022;227(5):737.e1-737.e11. doi: 10.1016/j.ajog.2022.06.054

29.	Rausch M, Lorch S, Chung K, Frederick M, Zhang J, Barnhart K. A cost-effectiveness analysis of surgical versus medical management of early pregnancy loss. *Fertil Steril* 2012;97(2):355-60. doi: 10.1016/j.fertnstert.2011.11.044

30.	Waitzman NJ, Jalali A, Grosse SD. Preterm birth lifetime costs in the United States in 2016: An update. *Semin Perinatol*. 2021;45(3). doi:10.1016/J.SEMPERI.2021.151390

31.	Rae M, Cox C, Dingel H. Health costs associated with pregnancy, childbirth, and postpartum care - Peterson-KFF Health System Tracker. Published July 13, 2022. Accessed October 23, 2023. https://www.healthsystemtracker.org/brief/health-costs-associated-with-pregnancy-childbirth-and-postpartum-care/

32.	Nissenson AR, Wade S, Goodnough T, Knight K, Dubois RW. Economic burden of anemia in an insured population. *J Manag Care Pharm*. 2005;11(7):565-574. doi:10.18553/JMCP.2005.11.7.565

33.	Soliman AM, Surrey E, Bonafede M, Nelson JK, Castelli-Haley J. Real-World Evaluation of Direct and Indirect Economic Burden Among Endometriosis Patients in the United States. *Adv Ther* 2018;35(3):408-423. doi: 10.1007/s12325-018-0667-3

34.	Yeh JM, Hook EW, Goldie SJ. A refined estimate of the average lifetime cost of pelvic inflammatory disease. *Sex Transm Dis*. 2003;30(5):369-378. doi:10.1097/00007435-200305000-00001

35.	Wang A, Wang S, Owens CD, Vora JB, Diamond MP. Health Care Costs and Treatment Patterns Associated with Uterine Fibroids and Heavy Menstrual Bleeding: A Claims Analysis. *J Womens Health (Larchmt)* 2022;31(6):856-863. doi: 10.1089/jwh.2020.8983

36.	Fink JL. How Common Are Miscarriages? | Miscarriage Rates, Statistics & Risk. Healthgrades. Published December 15, 2020. Accessed October 23, 2023. https://www.healthgrades.com/right-care/pregnancy/how-common-is-miscarriage

37.	Osterman MJK, Hamilton BE, Martin JA, Driscoll AK, Valenzuela CP. *National Vital Statistics Reports Volume 72, Number 1 January 31, 2023*.; 2021. https://www.cdc.gov/nchs/products/index.htm.

38.	Fennessy FM, Kong CY, Tempany CM, Swan JS. Quality-of-life assessment of fibroid treatment options and outcomes. *Radiology*. 2011;259(3):785-792. doi:10.1148/RADIOL.11100704/-/DC1

39.	O’Sullivan AK, Thompson D, Chu P, Lee DW, Stewart EA, Weinstein MC. Cost-effectiveness of magnetic resonance guided focused ultrasound for the treatment of uterine fibroids. *Int J Technol Assess Health Care*. 2009;25(1):14-25. doi:10.1017/S0266462309090035
