## Supplementary material for "Cost-effectiveness of Ultrasound Screening for Uterine Fibroids in the United States": Decision Tree and Assumptions

### **Appendix C: Decision Tree and Assumptions**

**Figure C.1. The Decision Tree Diagram Used in TreeAge Pro**

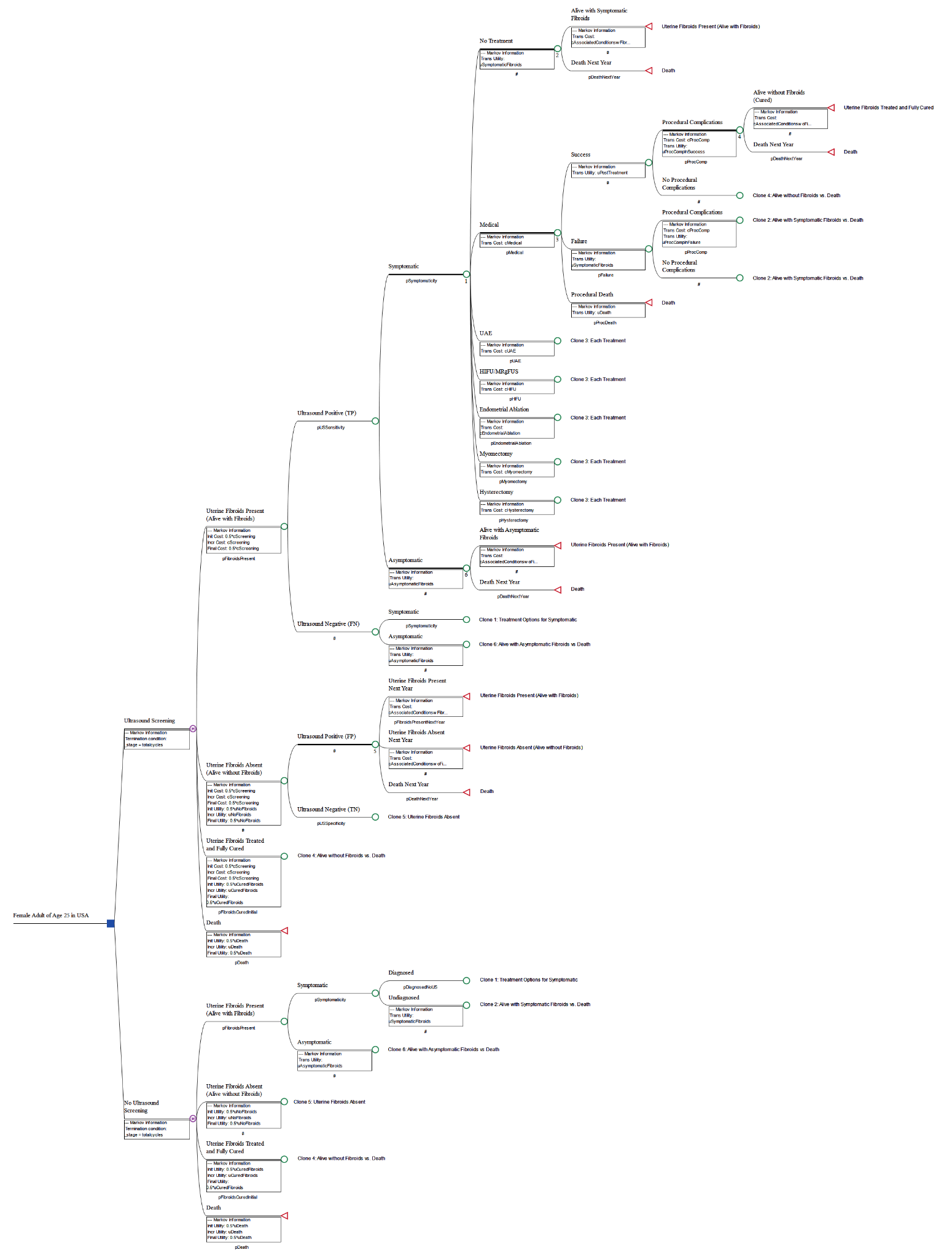

**Figure C.1 Legend:** Square node is a ‘Decision Node’ representing two options to be compared. The nodes, marked with the letter ‘M’, stand for Markov nodes. The triangular nodes are terminal nodes, which represent the end of the respective cycle, where transitions between health states occur (‘jumps’ back to one of the branches of the Markov node). ‘#’ indicates complementary probability to the other branches, so as to sum up to a total probability of 1.0 (100%). ‘Clone’ means that the node had similar further branches like the Master Node (numbered); where the probabilities and pay-offs (costs and utilities) might be same or different as applicable.

#### Simple Language Explanation of Decision Tree

When following the nodes of the Decision Tree, a woman could either receive ultrasound screening or no ultrasound screening at age 25. All women of younger ages might have not been comfortable with being examined using transvaginal ultrasound. Hence, we assumed the use of either transabdominal or transvaginal techniques for ultrasound screening of fibroids, as per the woman’s choice. In the Markov model, women following the decision arm of ultrasound screening stayed in the ultrasound screening arm (continue re-screening according to schedule). Similarly, women following the decision arm of no ultrasound screening stayed in the same arm, and thereby did not receive screening until the time they were clinically diagnosed.

Throughout the Markov Model, a woman could transition between four health states: Alive without Fibroids, Alive with Fibroids (Symptomatic or Asymptomatic), Alive without Fibroids (Successfully Treated), and Death. If the woman received annual ultrasound screening and it was true-negative, meaning no fibroids are present, then they would continue through the model in the “Alive without Fibroids” health state until they happened to develop fibroids in a future cycle. However, if the screening was true-positive and the fibroids are symptomatic, the woman could choose between the available medical, surgical, or no treatment options. The probability of the woman choosing each line of treatment depended on her age due to multiple factors, such as the desire for future fertility or the time spent suffering with symptoms.^1,2^ For each treatment option, there were possibilities of adequate recurrence-free relief (success), failure (including recurrence), or procedural death. If the treatment was a success, then the woman would continue through the model in the “Alive without Fibroids (Successfully Treated)” health state. On the other hand, if the treatment was a failure, or the woman initially chose to pursue no treatment, then they would continue in the “Alive with Fibroids (Symptomatic)” health state at the beginning of the next cycle. If the woman had asymptomatic fibroids, they would receive no treatment and will remain in the “Alive with Fibroids (Asymptomatic)” health state until symptoms happen to develop in a future cycle. If the ultrasound screening was false-negative, they would only be treated if symptomatic and clinically diagnosed. Additionally, we assumed that all false-positive cases (ultrasound-positive cases without fibroids) are asymptomatic. Even if these women had symptoms, owing to the definition of false-positive, the symptoms would be due to some other condition; hence, it would be appropriate to ignore that from the analysis. The fourth health state a woman could transition to was the “Death” health state, either due to procedural or background death (the statistical probability of death due to any reason for a particular age group as outlined by CDC reports).^3^

For each health state a woman could be in at the beginning of every cycle, there are associated costs and utility scores of that health state. Most significantly, each additional year a woman lived with symptomatic fibroids incurred costs of conditions associated with fibroids. A woman with symptomatic fibroids had a higher risk of spontaneous abortion, preterm delivery, cesarean delivery, anemia, endometriosis, inflammatory diseases, and menstruation disorders, as they are either results of the symptoms or vice versa.^1,4^ By taking the product of the risk of these conditions when symptomatic fibroids are present and their respective costs, this was the average cost that was accounted for when a woman continued to live with symptomatic fibroids. Since women could have these conditions even when fibroids were asymptomatic or absent, the product of the general risk of each condition and their respective costs were accounted for when a woman continued to live without symptomatic fibroids. However, the risk of having these conditions has been known to be much higher with symptomatic fibroids, leading to a greater associated cost the longer the woman’s symptomatic fibroids were present. While symptomatic fibroids have been shown to be associated with many more conditions, we have limited to the above conditions for the simplicity of the model and to maintain a conservative approach.

On top of costs that are associated with each health state, other costs such as the direct cost of an ultrasound screening and each treatment option were accounted for. While most obstetrician-gynecologists in the USA have facilities of outpatient ultrasonography in their clinics and are trained for the same (as required for board certification as stated by ABOG^5^); regular use of ultrasonography in well-woman clinic/office practice has been limited. Hence, we have attributed the costs of ultrasonography separately as an imaging center-based investigation. Here, we also assume that the sonographer involved is knowledgeable of what to look for, and competent to interpret the ultrasonographic findings, as most obstetrician-gynecologists and radiologists in the USA are trained in ultrasonography (required for board certification by ABOG and ABR). Additionally, for any of the surgical treatment choices, there could be a risk of procedural complications, regardless if the procedure was a success or failure, which also have added costs. Procedural complication rates included both minor and major complications, as well as the need for blood transfusions.

If a woman did not receive annual ultrasound screening for fibroids, which is the current standard practice, then they would only be treated if they are diagnosed with symptomatic fibroids. However, as mentioned earlier, the majority of women with fibroids went undiagnosed. This would cause many more women to remain in the “Alive with Fibroids (Symptomatic)” health state for longer, impacting the added costs and utility scores.

**References to Appendix C**

1.	Eltoukhi HM, Modi MN, Weston M, Armstrong AY, Stewart EA. The health disparities of uterine fibroid tumors for African American women: a public health issue. *Am J Obstet Gynecol*. 2014;210(3):194-199. doi:10.1016/j.ajog.2013.08.008

2.	Cain-Nielsen AH, Moriarty JP, Stewart EA, Borah BJ. Cost-effectiveness of uterine-preserving procedures for the treatment of uterine fibroid symptoms in the USA. *J Comp Eff Res*. 2014;3(5):503-514. doi:10.2217/CER.14.32

3.	Xu J, Murphy S, Kochanek K, Arias E. *Mortality in the United States, 2021*.; 2022. doi:10.15620/cdc:122516

4.	Cardozo ER, Clark AD, Banks NK, Henne MB, Stegmann BJ, Segars JH. The estimated annual cost of uterine leiomyomata in the United States. *Am J Obstet Gynecol*. 2012;206(3):211.e1-211.e9. doi:10.1016/J.AJOG.2011.12.002

5.	ABOG. Specialty Certifying Exam Preparation. Accessed October 23, 2023. https://www.abog.org/specialty-certification/certifying-exam/exam-preparation
