## Supplementary material for "Cost-effectiveness of Ultrasound Screening for Uterine Fibroids in the United States": Results of Alternative Analysis of Starting Age

### **Appendix D: Results of Alternative Analysis of Starting Age**

**Table D.1.** Effect of Starting Age of Screening on Cost-Effectiveness for All Women

| **Starting Age (Years)** | **Incremental Cost (US $ added)*** | **Incremental Effectiveness (QALYs gained)** | **ICER (US $ per QALY gained)*** | **Incremental NMB (US $)**  **[WTP: 30000]** |
| --- | --- | --- | --- | --- |
| **25** | -17,762.261 | 0.318 | -55,831.777 | 27,306.429 |
| **26** | -17,587.892 | 0.317 | -55,499.026 | 27,095.027 |
| **27** | -17,754.463 | 0.313 | -56,717.392 | 27,145.479 |
| **28** | -18,179.623 | 0.310 | -58,556.780 | 27,493.466 |
| **29** | -18,424.759 | 0.318 | -58,006.379 | 27,953.759 |
| **30** | -18,908.410 | 0.304 | -62,269.111 | 28,018.100 |
| **31** | -18,462.749 | 0.299 | -61,719.573 | 27,436.928 |
| **32** | -18,059.545 | 0.302 | -59,867.219 | 27,109.345 |
| **33** | -17,972.816 | 0.293 | -61,310.729 | 26,767.108 |
| **34** | -18,075.785 | 0.296 | -61,081.109 | 26,953.711 |
| **35** | -18,315.566 | 0.286 | -63,958.982 | 26,906.493 |
| **36** | -17,673.761 | 0.274 | -64,609.202 | 25,880.221 |
| **37** | -17,135.906 | 0.265 | -64,708.677 | 25,080.391 |
| **38** | -16,342.227 | 0.256 | -63,836.834 | 24,022.225 |
| **39** | -15,468.091 | 0.244 | -63,426.224 | 22,784.351 |
| **40** | -15,124.296 | 0.236 | -64,126.680 | 22,199.805 |
| **41** | -14,810.470 | 0.231 | -64,216.923 | 21,729.427 |
| **42** | -13,828.305 | 0.226 | -61,307.217 | 20,595.032 |
| **43** | -12,871.678 | 0.235 | -54,762.882 | 19,922.993 |
| **44** | -11,893.602 | 0.231 | -51,527.488 | 18,818.218 |
| **45** | -10,791.261 | 0.228 | -47,358.649 | 17,627.137 |
| **46** | -9,733.733 | 0.226 | -43,124.198 | 16,505.152 |
| **47** | -8,181.777 | 0.223 | -36,652.388 | 14,878.566 |
| **48** | -6,789.302 | 0.221 | -30,727.416 | 13,417.880 |
| **49** | -5,153.473 | 0.212 | -24,346.456 | 11,503.646 |
| **50** | -3,708.804 | 0.198 | -18,689.079 | 9,662.235 |
| **51** | -2,241.089 | 0.177 | -12,659.123 | 7,552.093 |
| **52** | -1,048.841 | 0.148 | -7,104.488 | 5,477.764 |
| **53** | -155.400 | 0.106 | -1,467.819 | 3,331.543 |
| **54** | 344.649 | 0.051 | 6,717.804 | 1,194.465 |

Abbreviations: QALY = Quality-Adjusted Life Years, ICER = Incremental Cost-Effectiveness Ratio, NMB = Net Monetary Benefit, WTP = Willingness-To-Pay Threshold

All values are rounded to the nearest three decimal places

*Negative values indicate cost-saving

**Table D.2.** **Effect of Starting Age of Screening on Cost-Effectiveness for Black Women**

| **Starting Age (Years)** | **Incremental Cost (US $ added)*** | **Incremental Effectiveness (QALYs gained)** | **ICER (US $ per QALY gained)*** | **Incremental NMB (US $)**  **[WTP: 30000]** |
| --- | --- | --- | --- | --- |
| **25** | -19,536.454 | 0.311 | -62,877.445 | 28,857.659 |
| **26** | -19,552.437 | 0.300 | -65,161.360 | 28,554.291 |
| **27** | -19,545.805 | 0.293 | -66,629.377 | 28,346.340 |
| **28** | -19,413.469 | 0.296 | -65,695.666 | 28,278.652 |
| **29** | -19,617.573 | 0.287 | -68,304.822 | 28,233.761 |
| **30** | -19,611.799 | 0.293 | -66,859.801 | 28,411.615 |
| **31** | -20,114.287 | 0.297 | -67,802.179 | 29,014.128 |
| **32** | -19,719.530 | 0.276 | -71,397.932 | 28,005.287 |
| **33** | -19,481.820 | 0.272 | -71,611.472 | 27,643.286 |
| **34** | -19,171.357 | 0.265 | -72,350.675 | 27,120.705 |
| **35** | -19,152.367 | 0.234 | -82,018.552 | 26,157.746 |
| **36** | -19,303.399 | 0.246 | -78,624.702 | 26,668.793 |
| **37** | -18,515.500 | 0.235 | -78,693.332 | 25,574.103 |
| **38** | -17,827.637 | 0.231 | -77,312.357 | 24,745.407 |
| **39** | -17,094.210 | 0.219 | -78,193.813 | 23,652.611 |
| **40** | -16,733.759 | 0.212 | -79,018.187 | 23,086.888 |
| **41** | -16,255.887 | 0.211 | -76,918.530 | 22,596.057 |
| **42** | -15,209.795 | 0.208 | -72,980.949 | 21,462.027 |
| **43** | -14,190.774 | 0.203 | -69,903.921 | 20,280.894 |
| **44** | -13,309.797 | 0.199 | -67,003.611 | 19,269.086 |
| **45** | -12,101.940 | 0.202 | -59,769.606 | 18,176.234 |
| **46** | -10,904.964 | 0.215 | -50,705.088 | 17,356.958 |
| **47** | -9,384.364 | 0.218 | -43,138.307 | 15,910.604 |
| **48** | -7,910.936 | 0.218 | -36,341.531 | 14,441.428 |
| **49** | -6,335.397 | 0.215 | -29,449.607 | 12,789.197 |
| **50** | -4,615.588 | 0.205 | -22,554.764 | 10,754.764 |
| **51** | -3,091.142 | 0.187 | -16,560.824 | 8,690.758 |
| **52** | -1,779.261 | 0.161 | -11,030.050 | 6,618.571 |
| **53** | -589.880 | 0.119 | -4,937.244 | 4,174.149 |
| **54** | -22.768 | 0.062 | -367.521 | 1,881.275 |

Abbreviations: QALY = Quality-Adjusted Life Years, ICER = Incremental Cost-Effectiveness Ratio, NMB = Net Monetary Benefit, WTP = Willingness-To-Pay Threshold

All values are rounded to the nearest three decimal places

* Negative values indicate cost-saving
