## Supplementary material for "Cost-effectiveness of Ultrasound Screening for Uterine Fibroids in the United States": Calculations of Cumulative Lifetime Economic and Health Outcomes

### **Appendix E: Calculations of Cumulative Lifetime Economic and Health Outcomes**

The female (all women) and black female populations were derived from the US Census, which listed the breakdown of population by age, sex, and race.^1,2^

**Table E.1.** **Female Population by Age in the US^1^**

| **Age range** | **Female Population (in thousands)** |
| --- | --- |
| 25 to 29 years | 11,345 |
| 30 to 34 years | 11,132 |
| 35 to 39 years | 10,840 |
| 40 to 44 years | 10,107 |
| 45 to 49 years | 10,147 |
| 50 to 54 years | 10,314 |
| Total | 63,885 |

**Table E.2.** **Black Female Population by Age in the US^2^**

| **Age range** | **Black Female Population (in thousands)** |
| --- | --- |
| 25 to 29 years | 1,844 |
| 30 to 34 years | 1,626 |
| 35 to 44 years | 2,972 |
| 45 to 54 years | 2,873 |
| Total | 9,315 |

In order to calculate the total amount of money and QALYs saved every year from performing ultrasound screening as compared to no ultrasound screening, we took the product of the female population in the US (for all women and black women) and the incremental cost or incremental effectiveness for that population, taken from base-case results.

**Table E.3.** **Total Costs Saved By Performing Ultrasound Screening for All Women and Black Women Populations**

| **Population** | **Incremental Cost** | **Calculation** | **Total Costs Saved** | **Total Costs Saved (rounded)** |
| --- | --- | --- | --- | --- |
| All women | - $25,562.330* | 63,885,000 x 18,301.720 | $1,169,205,382,200 | ~ $1,169 billion |
| Black women | - $26,956.600* | 9,315,000 x 19,683.438 | $183,351,224,970 | ~ $183 billion |

**Table E.4. Total QALYs Gained From Performing Ultrasound Screening for All Women and Black Women Populations**

| **Population** | **Incremental Effectiveness** | **Calculation** | **Total QALYs Gained** | **Total QALYs Gained (rounded)** |
| --- | --- | --- | --- | --- |
| All women | 0.323 | 63,885,000 x 0.323 | 20,655,284 | ~ 20.7 million |
| Black women | 0.323 | 9,315,000 x 0.323 | 3,008,971 | ~ 3 million |

**References to Appendix E**

1.	Age and Sex Composition in the United States: 2020. US Census Bureau. Published 2020. Accessed October 23, 2023. https://www.census.gov/data/tables/2020/demo/age-and-sex/2020-age-sex-composition.html

2.	The Black Alone Population in the United States: 2019. US Census Bureau. Published 2019. Accessed October 23, 2023. <https://www.census.gov/data/tables/2019/demo/race/ppl-ba19.html>
